# Long-read sequencing resolves the clinically relevant *CYP21A2* locus, supporting a new clinical test for Congenital Adrenal Hyperplasia

**DOI:** 10.1101/2025.02.07.25321404

**Authors:** Jean Monlong, Xiao Chen, Hayk Barseghyan, William J Rowell, Shloka Negi, Natalie Nokoff, Lauren Mohnach, Josephine Hirsch, Courtney Finlayson, Catherine E. Keegan, Miguel Almalvez, Seth I. Berger, Ivan de Dios, Brandy McNulty, Alex Robertson, Karen H. Miga, Phyllis W. Speiser, Benedict Paten, Eric Vilain, Emmanuèle C. Délot

## Abstract

Congenital Adrenal Hyperplasia (CAH), one of the most common inherited disorders, is caused by defects in adrenal steroidogenesis. It is potentially lethal if untreated and is associated with multiple comorbidities, including fertility issues, obesity, insulin resistance, and dyslipidemia. CAH can result from variants in multiple genes, but the most frequent cause is deletions and conversions in the segmentally duplicated RCCX module, which contains the *CYP21A2* gene and a pseudogene.

The molecular genetic test to identify pathogenic alleles is cumbersome, incomplete, and available from a limited number of laboratories. It requires testing parents for accurate interpretation, leading to healthcare inequity. Less severe forms are frequently misdiagnosed, and phenotype/genotype correlations incompletely understood. We explored whether emerging technologies could be leveraged to identify all pathogenic alleles of CAH, including phasing in proband-only cases. We targeted long-read sequencing outputs that would be practical in a clinical laboratory setting.

Both HiFi-based and nanopore-based whole-genome long-read sequencing datasets could be mined to accurately identify pathogenic single-nucleotide variants, full gene deletions, fusions creating non-functional hybrids between the gene and pseudogene (“30-kb deletion”), as well as count the number of RCCX modules and phase the resulting multimodular haplotypes. On the Hi-Fi data set of 6 samples, the PacBio Paraphase tool was able to distinguish nine different mono-, bi-, and tri-modular haplotypes, as well as the 30-kb and whole gene deletions. To do the same on the ONT-Nanopore dataset, we designed a tool, Parakit, which creates an enriched local pangenome to represent known haplotype assemblies and map ClinVar pathogenic variants and fusions onto them. With few labels in the region, optical genome mapping was not able to reliably resolve module counts or fusions, although designing a tool to mine the dataset specifically for this region may allow doing so in the future.

Both sequencing techniques yielded congruent results, matching clinically identified variants, and offered additional information above the clinical test, including phasing, count of RCCX modules, and status of the other module genes, all of which may be of clinical relevance. Thus long-read sequencing could be used to identify variants causing multiple forms of CAH in a single test.

## Introduction

With an incidence rate of 1 in 10,000 to 18,000 live births, congenital adrenal hyperplasia (CAH) is one of the most common inherited disorders. CAH can be caused by multiple genes but the most frequent etiology (∼95% of cases, OMIM #201910) is a recessive condition where impaired 21-hydroxylase activity blocks conversion of the primary ligand, 17-hydroxyprogesterone (17-OHP) to 11-deoxycortisol in the adrenal cortex, resulting in reduction of cortisol, and in severe cases, aldosterone, with concomitant excessive production of androgens. The severity of 21-hydroxylase deficiency phenotype is linked to the degree of residual enzyme activity. Phenotypes range from potentially lethal classic salt-wasting (SW-CAH, in ∼75% of affected newborns), to non-lethal virilizing, to later-onset non-classic form. 46,XX infants with classic CAH caused by null *CYP21A2* variants are born with genital atypia (including clitoromegaly, labial fusion, urogenital sinus), whereas 46,XY individuals have typical male genitalia. Non-classic CAH is less severe but more frequent, with prevalence estimates of 1/200 in the US population, possibly higher in Hispanics and other ethnic groups (Speiser, 1985; Hannah-Schmouni 2017; Livadas 2020; Jha 2021). Both XX and XY individuals may be affected but the non-classic phenotype is more typically diagnosed in 46,XX individuals, where it may manifest in mid-childhood with premature adrenarche or in adolescents with hirsutism, acne, and menstrual disturbances. Subfertility, obesity, insulin resistance, and dyslipidemia are frequently associated with all forms CAH (Claahsen van der Grinten 2022).

Because of its severity, relative frequency, and available treatment, CAH caused by CYP21A2 deficiency is assayed on most newborn screens by biochemical detection of 17-OHP concentrations. This screen has a highly variable positive predictive value depending on the protocol, reported as low as 0.7% (Speiser 2020), and a positive finding requires confirmation. However, *CYP21A2* genotyping is error-prone due to the complexity of the genomic region (reviewed in the 2020 best practice guidelines by the European Molecular Genetics Quality Network, Baumgartner-Partzer 2020). The molecular genetic test to identify pathogenic alleles is cumbersome, available from a limited number of laboratories, and frequently not offered to patients. This limits the ability to offer precise genetic counseling for family planning, and results in frequent misdiagnosis of less severe forms, and an incomplete understanding of phenotype/genotype correlations (for a comprehensive summary of CAH etiology, phenotypes, and clinical management, see Claahsen 2022).

### The CYP21A2 gene

The *CYP21A2* gene sits on the short arm of human chromosome 6, in the HLA Class III region (6p21.33) between the regions encoding the HLA Class I and Class II complexes. It is nestled inside a ∼30 kb module, termed the RCCX module (**Figure 1A**), with three other genes, two of which are also of clinical relevance, *TNXB* (where biallelic variants cause Ehlers-Danlos syndrome, classic-like, 1, OMIM #606408; Burch 1997; Schalkwijk 2001) and *C4* (associated with susceptibility to schizophrenia and autoimmune conditions; Kamitaki 2020). Most of the population carries bimodular alleles with the first (telomeric) of the modules containing a pseudogene, *CYP21A1P*, and the other a copy of the active *CYP21A2* gene. The pseudogene and the gene are 98 and 96% identical in the coding and non-coding regions, respectively, with the pseudogene carrying multiple recurrent mutations and a weak promoter. Similarly, copies of the *C4* gene sit in tandem duplication upstream of the gene and pseudogene. The *TNXB* gene and its truncated pseudogene *TNXA* are transcribed from the opposite strand and overlap with the 5’UTRs of *CYP21*. The dizzying complexity of the region is completed by the presence, in each module, of an adrenal enhancer for *CYP21A2* inside the *C4* gene, a non-coding RNA overlapping with both CYP21- and TNX-encoding genes, and a polymorphic 6.4 kb retroviral transposon (HERV-K) in exon 9 of *C4* deleted in 40% of the population in the second, but not the first module (for further details on the complexity of the region, see Miller 2021).

**Figure 1:**
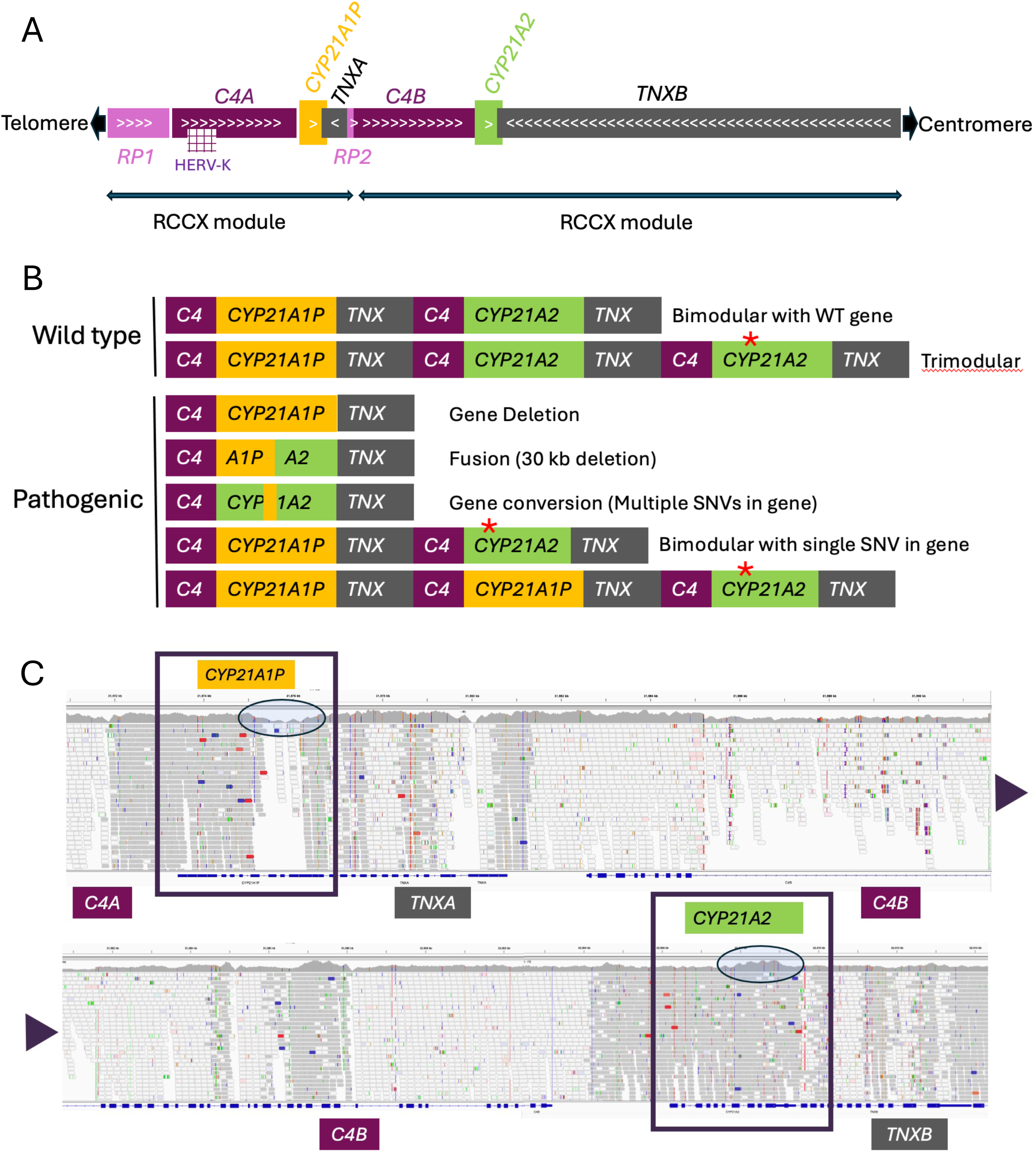
Structure of the RCCX module containing the *CYP21A2* gene. **A:** Organization of the locus in a bimodular allele. >> indicates gene orientation. The *CYP21* and *TNX* genes overlap and are encoded from opposite strands. The HERV-K transposon is shown in the *C4A* gene; it is frequently absent in the second module in the population. The same color conventions for *CYP21A1 (orange)*, *CYP21A2 (green)*, and *TNX (dark grey)* are used throughout the manuscript. **B:** Cartoons of frequent RCCX alleles (Colored blocks for the various genes of the module are not to scale of gene length; the *RP* genes are not represented). The most frequent in the population is a bimodular haplotype with one *CYP21A1P* pseudogene and one *CYP21A2* gene. A frequent allele is trimodular with one pseudogene and two genes, one of them carrying the Q319X pathogenic variant (red asterisk). As CAH is a recessive condition, this is a wild type allele. Pathogenic haplotypes include whole gene deletion (resulting in CAH-X syndrome is the fusion occurs in *TNXB*), creation of an inactive hybrid 5’-*CYP21A1P/CYP21A2*-3’ fusion by deletion of the ∼30 kb module, gene conversions exchanging small pieces of the pseudogene and the gene, and bi- or tri-modular alleles with a variant in the only copy of the active gene. **C:** IGV visualization of Illumina short-read sequencing of the RCCX region. Reads in the region of the *CYP21A2* and *CYP21A1* pseudogene highlighted in the blue oval are all misaligned to the gene by algorithms. *C4A/C4B* and *TNXA/TNXB* are also mostly unresolved.

### Identifying the variants causing CYP21A2 deficiency

As schematized in **Figure 1B**, pathogenic alleles resulting in CAH include (mostly recurrent) single nucleotide variants (SNVs), micro-conversions transferring pseudogene mutations to the active gene, and deletions leading to fusion events creating inactive *CYP21A1P/CYP21A2* chimeras or complete gene deletions (recently reviewed in Kim et al. 2023). Alleles are typically inherited from a parent. More than 200 discrete causative variants have been described. To assay these diverse events, the most comprehensive clinical test available includes 4 long-range PCRs across modules, bidirectional Sanger sequencing of the fragments that do amplify, and MLPA to count the number of genes and pseudogenes. Even then, alleles are not phased, and testing parents is critical to avoid misinterpretation. This carries the risk of healthcare inequity through increased cost and penalizing families where both biological parents are not available for testing. Predictably, the region has historically been intractable with short-read sequencing (**Figure 1C**).

### Module counts are clinically relevant

Many clinical laboratories typically do not offer this full test. Some detect only the frequent recurrent SNVs and the most frequent alleles (fusion and gene deletion) are not found. In the absence of phasing and module-counting, this can lead to interpretation errors. For example, a frequent (1-2%) allele in the general population is trimodular, with one pseudogene and two copies of the active gene, one of which is carrying the recurrent stop-gain variant p.Gln319Ter (Q319X) (**Figure 1B**). With one intact copy of *CYP21A2*, it is a wild-type allele, but the presence of Q319X could easily be misinterpreted as pathogenic in the absence of complete characterization and phasing.

In addition, a condition resulting from a continuous gene deletion involving both *TNXB* and *CYP21A2*, termed CAH-X syndrome, was reported to affect about 10% of people with CYP21A2-related CAH (Gao 2020). CAH-X carries the risks associated with hypermobility forms of Ehlers-Danlos syndrome, including cardiac structural defects, joint instability, hernias, and gastrointestinal disorders. While this was described decades ago (Burch 1997; Merke 2012), the phenotype is not routinely documented in the follow-up of patients, and current molecular testing for CAH does not determine *TNXB* status. CAH-X was shown to be particularly prevalent (30%) in patients carrying the most common CAH allele, the so-called 30-kb deletion resulting in a fusion between gene and pseudogene, and experts have recommended *TNXB* genotyping should be included in CAH testing (Lao 2021). Such a test would be particularly advantageous to identify at-risk children too young to be evaluated for hypermobility.

We tested whether long-read genome sequencing (LRS) and/or optical genome mapping (OGM) could provide a new, complete test for CYP21A2-related CAH. As standards for long-read sequencing are still emerging, we aimed to identify conditions (e.g. no ultralong reads, ∼30x coverage) and common tools to approximate what would be practical and economically viable in a clinical setting.

## Materials and Methods

High-molecular-weight DNA extracted from the blood of participants in the DSD-Translational Research Network (DSD-TRN; Délot 2017) was subjected to OGM (Bionano Genomics platform) and/or LRS on both the ONT Nanopore platform and the PacBio Revio instrument. Clinically detected variants were initially hidden to the OGM and LRS dataset analysts to test the abilities of existing algorithms.

### Participant samples

Patient samples sent to the DSD-TRN biobank from 4 sites of the network were used in this project. All but Proband #4 had previously undergone clinical genetic testing, in a variety of settings. In cases where the original report was not available to the clinical team, we used the language found in clinic notes to describe their pathogenic variants. All patients had the severe, salt-wasting, form of CAH. None had a comprehensive evaluation of phenotypic traits of CAH-X syndrome, but none had complaints related to hypermobility to date.

Summary of clinical test results and research tests performed for this study are shown for probands and family members in **Table 1**. (Further demographic, clinical, and genetic information for the probands are withheld per Medrxiv policy).

**Table 1:**
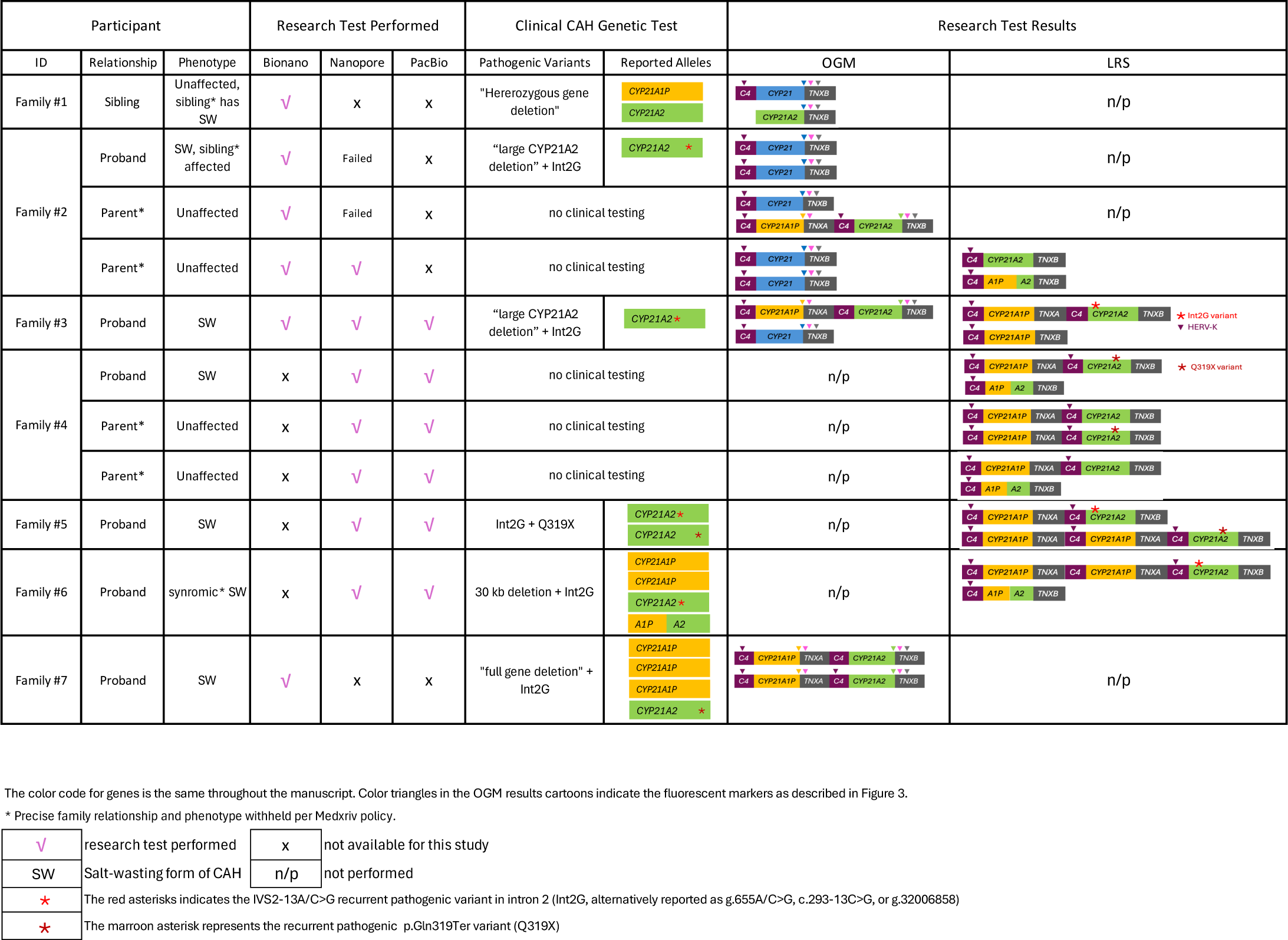
Summary of the cohort, research tests performed, and cartoons of the alleles identified with each technique.

> **Participant #1 (singleton):** The patient presented to Pediatric Endocrinology in mid-childhood due to rapid height and weight gain and body odor. Given her sibling’s diagnosis of classic, SW-CAH, there was a question whether she could have non-classic CAH. ACTH stimulation testing showed elevated 17-OHP and 17-OH pregnenolone, both of which could indicate CAH. She subsequently underwent full sequence analysis at Mayo Genetics Laboratory, which revealed one copy of *CYP21A1P* and a deletion of one copy of *CYP21A2*. No variants were found in the remaining active gene. She was counseled to be a carrier who did not require treatment for CAH.
>
> Clinical testing in her (affected) sibling had identified that both copies of the active *CYP21A2* genes were deleted, with one copy the likely inactive 5’CYP21AP/CYP21A2-3’ hybrid (representing the 30 kb deletion), and one copy of the pseudogene. The proband’s blood was not available for this study as it had been sent to the DSD-TRN biobank in 2013, before samples were preserved adequately for high-molecular weight (HMW) DNA extraction. Participant #1’s sample was processed as a proxy at a time she had a likely diagnosis of CAH and was thought to carry the same pathogenic alleles as the proband.
>
> **Proband #2 (trio):** In this patient, tested at birth because her sibling was known to have SW-CAH, full sequence analysis at Mayo Genetics Laboratories identified a gene deletion and the recurrent IVS2-13A/C>G splice acceptor site mutation in intron 2 (Int2G) in the remaining *CYP21A2* gene. Number of copies of pseudogene were not recorded in clinical notes and the original report was not found. Parents and sibling did not undergo clinical genetic testing but parents provided samples for this study.
>
> **Proband #3 (singleton):** This adolescent female with SW-CAH underwent testing abroad in infancy because of reported “genital ambiguity” at birth. She had genitoplasty at another center and notes prior to surgery are not available. Post-operatively, she was noted to have enlarged labia with some rugation, with clitoris and clitoral hood of a typical size. Variants were reported as Int2G and “large deletion” during intake with the US clinical team, but the original report was not available. She had surgery for rectal prolapse; it is unclear if this is a symptom of CAH-X.
>
> **Proband #4 (trio):** After a newborn screen positive result, a repeat measure of 17-OHP was also grossly elevated, consistent with classic CAH. Androstenedione, total testosterone, and ACTH were also elevated. Genital exam in the first month of life showed clitoromegaly, chordee with single perineal opening, fused labioscrotal folds, no palpable gonads, and an anteriorly displaced anus. Parents were not interested in further clinical testing for *CYP21A2*, and this trio was examined with LRS and OGM as undiagnosed.
>
> **Proband #5 (singleton)**: An enlarged clitoris and partial labial fusion were noted at birth. Newborn screen and confirmatory elevated 17-OHP concentrations were consistent with CAH. Pelvic ultrasound showed a uterus, urogenital sinus, and ovaries. Clinical testing at Prevention Genetics identified two pathogenic SNVs, Int2G and Q319X. The test, which includes sequencing of PCR products but no MLPA, does not provide information on the number of gene or pseudogene copies present and does not phase variants.
>
> **Proband #6 (singleton):** This child was born to a mother who had experienced multiple early pregnancy losses, with a meatus at the base of an enlarged clitorophallic structure, partly fused hyperpigmented labia majora, and additional syndromic traits (potentially identifying specifics withheld per medxriv policy). A VCUG at birth could not resolve the type of urethra, elongated female vs. male with hypospadias. A cosyntropin stimulation test showed no increase of cortisol, suggesting adrenal insufficiency. The newborn screen showed an elevated 17-OHP, indicative of CAH, which was confirmed by results of the cosyntropin stimulation also resulting in markedly elevated serum 17-OHP. Clinical testing at Mayo Genetics Laboratories reported that MLPA detected one copy of the *CYP21A2* gene, two copies of the inactive *CYP21A1P* pseudogene and one copy of an inactive *CYP21A1P/CYP21A2* hybrid (“30-kb deletion”). Sequence analysis of the *CYP21A2* gene detected the Int2G variant.
>
> **Proband #7 (singleton):** Full gene analysis was performed at Mayo Genetics Laboratories at birth to confirm a report of positive newborn screen. A hemizygous g.655A/C>G variant (Int2G) and a *CYP21A2* full gene deletion were reported. MLPA identified 3 copies of the pseudogene and a single copy of the active gene. The patient has salt-wasting CAH, and presented at birth with posterior labial fusion, a single urogenital opening, and an enlarged clitorophallic structure.

### Optical Genome Mapping

High molecular weight (HMW) DNA was extracted from fresh/frozen whole blood using the SP Blood and Cell Culture DNA Isolation Kit (Bionano, catalog #80042). Between 750 - 1000 ng of HMW DNA was labeled with DLE-1 enzyme using the Direct Label and Stain (DLS) Kit (Bionano, catalog #80005). Labeled DNA was imaged on a Bionano Saphyr G2 instrument using Saphyr G2.3 chips (Bionano, catalog #20366). Only molecules greater than 150 kbp in size were used for further analysis. Individual sample metrics are available in Supplementary **Table S2**.

Output was analyzed using Access software [Bionano Access v1.7.2 and Bionano Solve v3.5, 3.6, 3.7]. Analyses were performed using *de novo* assembly and rare variant analysis pipelines on both reference genome assemblies GRCh37 (hg19) and GRCh38 (hg38) as indicated. Filtering criteria were as follows: BED SV overlap precision: 3 kbp; BED CNV overlap precision: 15 kbp; SV masking filter: “all”; copy number type: “all”; copy number confidence: 0.99; copy number min size: 500 kbp; self molecule count: 5; % in control: 10; SV chimeric score: “pass”; found in self molecules: “yes”. SV confidence filters were 0 for insertion and deletion, 0.7 for inversion, -1 for duplication, 0.3 and 0.65 for intra- and inter-chromosome translocations respectively.

### Long-Read HiFi sequencing

DNA was extracted with the Prep SP Blood and Cell Culture DNA Isolation kit (Bionano Genomics, catalog #80042) on blood from 6 samples, an undiagnosed trio and three diagnosed singletons, as part of a larger cohort (n=98). Samples were prepared following the standard operating procedure “Preparing whole genome and metagenome libraries using SMRTbell® prep kit 3.0” (available at PACB.com) using 1.5-3 µg of DNA. Libraries were constructed on the Hamilton Microlab Star and VENUS 5 system. Genomic DNA quality, concentration, and post size-selected SMRTbell library samples were quantified using the Qubit DNA HS assay (Thermo Fisher Scientific) and FEMTO Pulse (Agilent Technologies). The libraries were loaded at an on-plate concentration of 90 pM for HiFi sequencing on a Revio instrument at Pacific Biosciences with Revio Polymerase Kit, following the Revio SMRTlink setup.

For the 6 samples, read length mean was 17,533 base pairs (bp) [range 15,737-18,932] similar to the rest of the cohort (mean ± standard deviation: 18,185 ± 2,610). Mean aligned coverage depth was 34.83x (32.95 ± 4.77 for the cohort). Individual sample metrics are in **Table S2**.

Sequence outputs were processed with PacBio WGS Variant Pipeline v1.1.0, which uses Phrank scores (Jagadeesh 2018) to rank genes based on their associations with phenotypes (Python implementation protocol available on GitHub). Phrank indicated that *CYP21A2* was highly associated with the CAH Human Phenotype Ontology term and the rare single nucleotide *CYP21A2* variants identified in these samples are reported below.

As *CYP21A2* was one of the original nine genes of clinical interest in segmental duplications to be resolved by the Paraphase algorithm (X. Chen 2023; X. Chen 2025), sequences were examined using Paraphase v2.2.3. The output mini-BAMs restricted to the region generated by Paraphase show the consecutive RCCX modules collapsed and in order from 5’ to 3’, with the reads of the two haplotypes labeled in different colors. The six samples reported here are also part of the cohort used to describe the continued development of Paraphase (X. Chen 2025; corresponding study IDs are shown in **Table S2**).

### Long-read nanopore-based sequencing

Nine samples (the same six as with HiFi, plus an additional trio) were submitted for sequencing at UCSC as part of a larger cohort of 98 samples. Four samples were submitted as frozen whole blood, four as HMW DNA previously extracted for OGM, and one as frozen white blood cell pellet (Supplementary **Table S2**). Two of the additional trio, including the proband, failed extraction; the third was also of poor quality with the lowest yield of the entire cohort by far (Family #2 Mother, reported as Sample DSDTRN19 in Negi, 2025).

Details of the protocol can be found in Negi (2025). Briefly, HMW DNA was sheared to a peak size of approximately 50 kb. Sequencing was performed on a PromethION48 sequencer using R10.4.1 flow cells. Diploid *de novo* phased assemblies were generated using the Napu end-to-end pipeline (Kolmogorov 2023), and variant calls were harmonized against the GRCh38 reference genome. Yield for the seven successfully processed CAH samples averaged 96.8 Mbp [range: 58.6 to 133.9 Mbp], below the average of 111.6 Mbp for the whole cohort, due to the two samples processed from HMW DNA previously extracted for OGM. This represents an average coverage of 31.2x [range: 18.9-43.2], assuming a 3.1 Mbp genome. Individual sample metrics are in Supplementary **Table S2**.

### Parakit for identification, annotation and visualization of variants

We developed a method, *Parakit*, to call the variants in the RCCX region on ONT long reads. The reads are re-mapped to a pangenome with all the RCCX modules of each allele collapsed. In this pangenome graph, *nodes* correspond to nucleotide sequences, *edges* to adjacency in the input haplotypes, and *paths* to haplotypes. Below, we call module *P* the module containing the pseudogene *CYP21A1P*, and module *G* the other module containing the active gene *CYP21A2* **(Figure 2)**.

**Figure 2:**
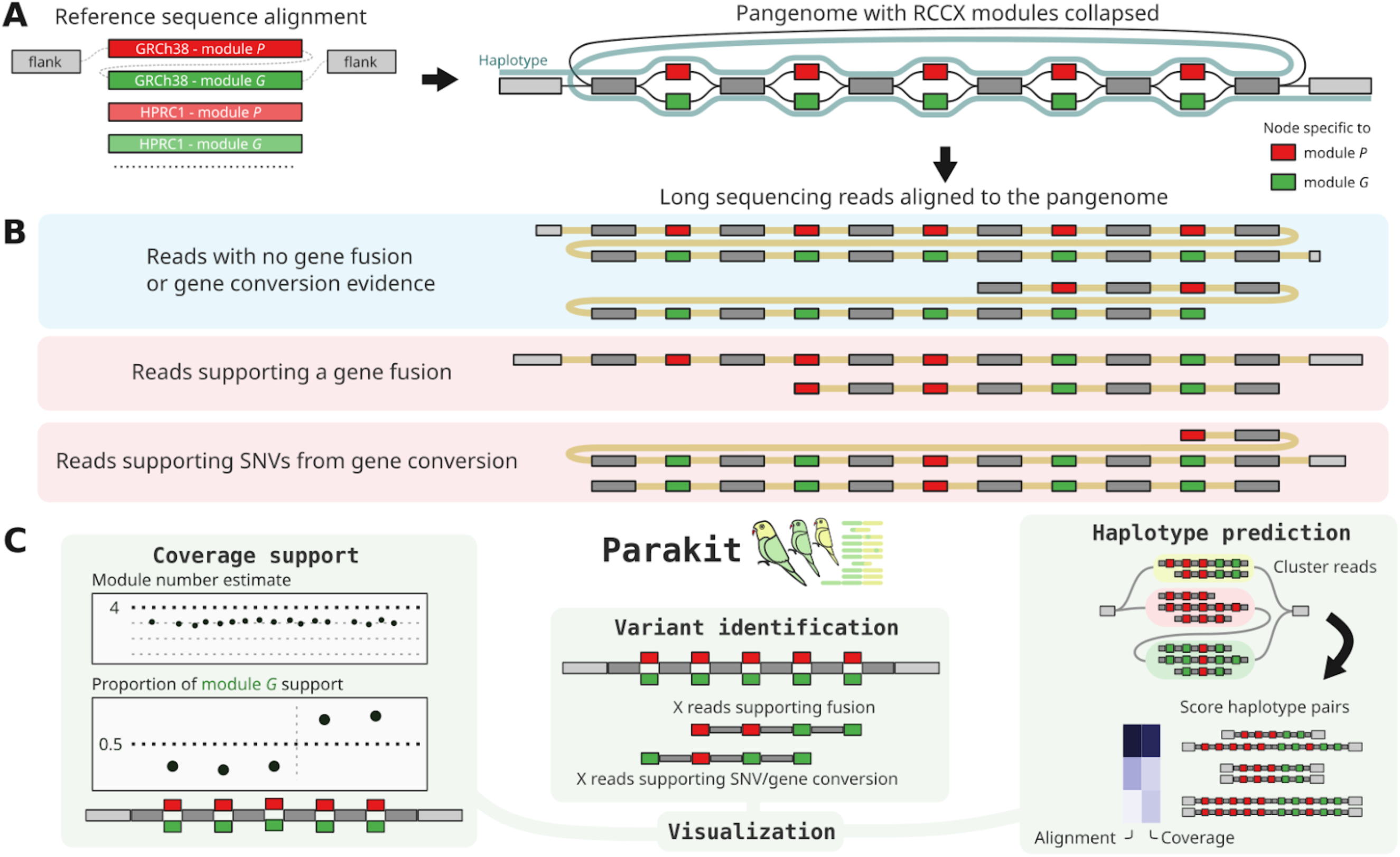
Identification, annotation and visualization of variants with Parakit. **A.** High-quality references are used to build a pangenome where the RCCX modules are collapsed. Nodes traversed mostly by module *P* or *G* are flagged and used to guide genome inference downstream. **B.** Long sequencing reads from a sample are aligned to the local pangenome. Based on the sequence of nodes traversed by each read, patterns of gene fusion or gene conversion can be detected, for example when a read switches from module *P* nodes to module *G* nodes. **C.** Parakit investigates the presence of variants from the aligned reads in three different analyses: using the coverage across nodes (left), aggregating variant support across reads (middle), reconstructing haplotypes (right). Parakit produces graphs combining the results of all three analyses.

*Local pangenome construction.* The pangenome is built using Minigraph-Cactus (Hickey 2024). The input sequences include first a modified version of the reference genome GRCh38 with module *G* removed. To this backbone, Minigraph-Cactus adds the module *G* from GRCh38, and module *P* and *G* from high-quality phased assemblies from the Human Pangenome Reference Consortium (HPRC, Liao 2023). For the latter, we selected the 65 haplotypes from the HPRC dataset that were consistently annotated as a bimodular allele by CAT (Fiddes 2018) and Ensembl. The full GRCh38 reference path is added to the pangenome using the *augment* subcommands of the vg toolkit (Garrison 2018). Finally, we identify nodes that tend to be specific to module *P* or *G* by counting the number of input modules from HPRC traversing them. For example, a node supported by three times more *P* input modules than *G* input modules is classified as *P*-specific. This classification is used for inference and visualization, as explained below.

*Long read re-alignment.* This pangenome, containing the sequence of 132 RCCX modules, is then used as a reference to re-align reads from this region, using GraphAligner (Rautiainen 2020). The reads map to only one position in this pangenomic reference because the RCCX modules are collapsed.

*Variant identification.* The aligned reads that traverse the pangenome through nodes that are specific to a module (as defined above) can be used for inference. *Parakit* uses this information to identify and visualize read signals supporting fusion or gene conversion events. First, read coverage helps estimate the total number of module copies. Parakit computes two estimates: the median read coverage normalized by the coverage in flanking regions and the read coverage around the nodes involved in the cycle (see Supplementary information). Second, changes in the allelic balance at informative sites, *i.e.* the proportion of reads traversing the nodes specific to module *P* compared to the ones specific to module *G*, help locate the breakpoints of a fusion or the boundaries of a large gene-converted region. Finally, *Parakit* searches for evidence of gene fusion or gene conversion in each read separately. Specifically, a sliding window approach looks for positions - within the RCCX module - where a read switches from aligning to *P*-specific nodes to aligning to *G*-specific nodes. The evidence is aggregated and reported if supported by at least 3 reads. SNVs predicted from gene conversions are also matched with the ClinVar database (Landrum 2014) to identify SNVs known to be pathogenic or likely pathogenic.

*Haplotype inference.* Parakit also infers the most likely pair of haplotypes by finding the pair of paths through the pangenome graph that are the most consistent with the aligned reads. First, haplotype candidates are enumerated. Reads are split at the module level and clustered into module consensus candidates. These module candidates are then combined into all possible haplotypes suggested by read junctions. The best pair of walks is then selected as the one maximizing read alignment scores and the correlation between observed coverage and coverage across the candidate pair of haplotypes. In summary, Parakit finds the pair of predicted haplotypes whose sequence and copy number best match the sequenced reads.

*Visualization.* Parakit produces graphs displaying the different layers of evidence described above on the collapsed pangenome. It includes a gene annotation track with the exon and intron for the genes in the region. Phasing information, for example to identify compound heterozygous variants, is suggested by mutually exclusive reads supporting the variants and predicted by the haplotype pair inferred.

*Availability.* Parakit is publicly available on GitHub. The RCCX pangenome and pipeline to perform the analysis are included in the repository.

## Results

### Optical Genome Mapping

While OGM does not allow base-level resolution, and would not be able to identify pathogenic SNVs, it is a relatively inexpensive method to identify large copy number variants and other structural variants (Barseghyan 2017; Mantere 2021; Barseghyan 2024). We tested whether it could replace the current cumbersome clinical method to identify the common 30-kb deletion and whole gene deletion alleles.

In the region of interest, each module had a very limited number of fluorescent markers. We highlighted each with triangles of different colors to facilitate interpretation of the output of the Access software (see cartoon under hg38 reference tracks in **Figure 3A**). One marker is in the HERV-K retroviral transposon (purple triangle); when absent, it was accurately called as a 6.5 kb deletion. One label is in exon 8 (see Supplementary **Figure S1**) of the *CYP21* pseudogene (orange) and gene (green), and one is common to *TNXA* and *TNXB* (pink). Beyond that, one marker is specific to *TNXB*, the longer version of the gene (grey). The reference (both hg19 and hg38) thus shows a pattern of 3 labels in the first RCCX module and 4 in the second, which allows ordering of the modules if all labels are present.

**Figure 3.**
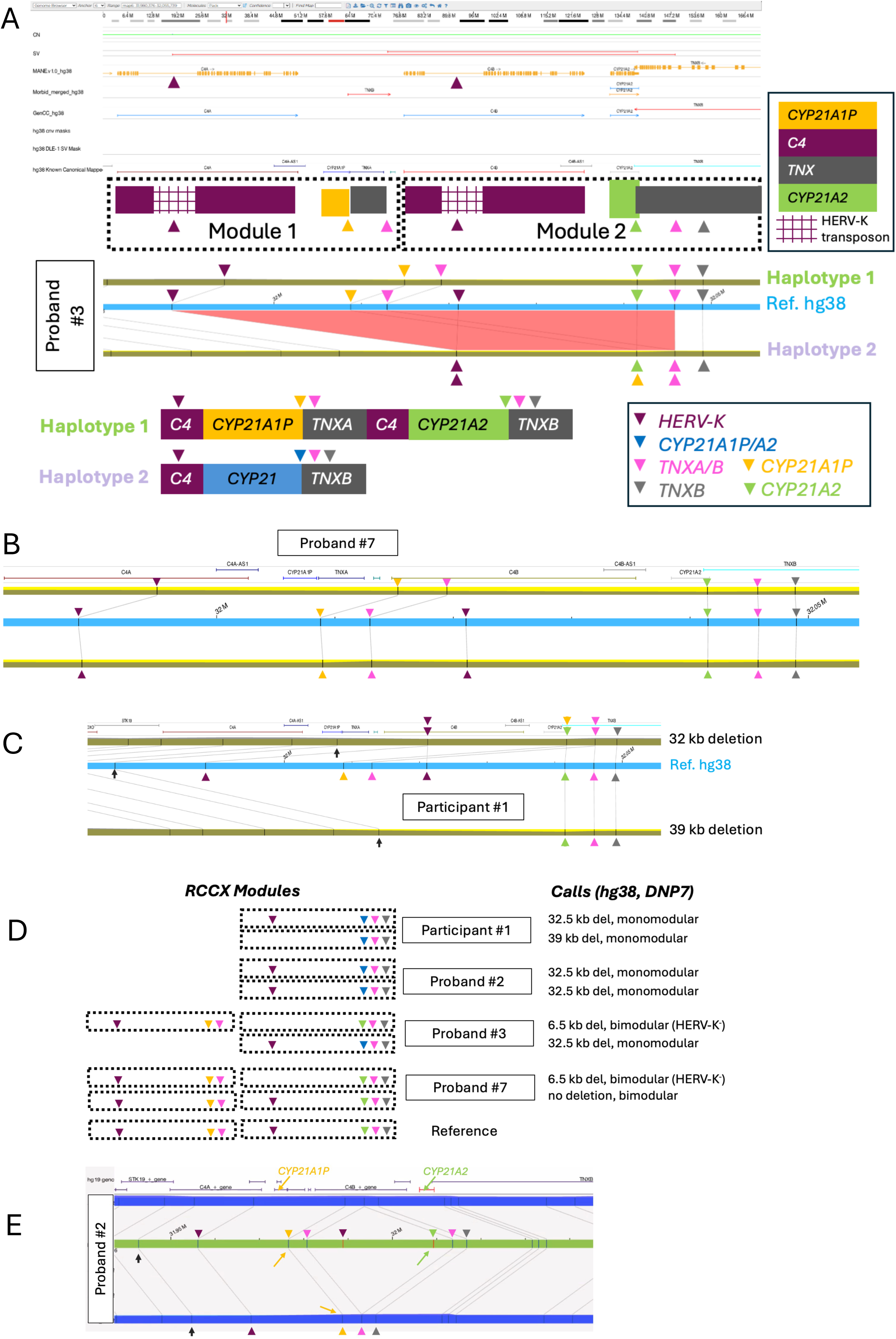
Optical genome mapping struggled to resolve deletion and poly-modular alleles. **A**: OGM detected a deletion in Proband #3. The three clinical genes of the RCCX module are schematized under the hg38 gene map of the Access software output, with the color code indicated in the inset at right and the modules framed with a dotted black line. The two haplotypes of the patient are shown in olive green above and below the reference assembly (in blue). DLE-1 labels are output as black vertical bars and we further identified each with different color triangles. The purple triangle is in the HERV-K retrovirus, in intron 9 of the *C4* gene. The grey triangle indicates a label specific to the *TNXB* gene. The label common to *TNXB* and *TNXA* is indicated with a pink triangle. The labels at the ends of the *CYP21A2* gene and the *CYP21A1P* pseudogene are shown with green and orange triangles respectively (see Supplemental **Figure S1**). Haplotype 1 appeared bimodular with HERV-K in the first, but not the second, RCCX module. This was (appropriately) called as a ∼6.5 kb deletion, which was filtered out because of high population frequency. The labels at the ends of *CYP21A2* and the pseudogene were both present. In Haplotype 2, a deletion was called by the Access software (visualized as a guava-colored irregular quadrangle between the proband and reference assemblies) with one HERV-K and the *TNXB*-specific labels preserved. The software mapped the two “pink” labels of the reference to a single label in the proband. Similarly, the reference green and yellow labels mapped to the same label, suggesting a monomodular haplotype with deletion of either *CYP21A2* or *CYP21A1P*. The haplotypes as inferred from the software output are represented below with the same color convention as in Figure 1. Blue color was used to indicate when *CYP21A2* and *CYP21A1P* could not be distinguished. **B**: In proband #7, OGM software output showed two bimodular haplotypes (olive green). The bottom one appeared identical to the hg38 reference assembly (blue). The top one appeared identical to Hap1 in Proband#3, with labels for both gene and pseudogene present and absence of HERV-K (purple triangles) in the 2^nd^ module. In both haplotypes of Proband #7, the (green) label at the end of the active *CYP21A2* gene was present. The three pseudogenes and full gene deletion detected clinically in this patient were not visualized in the OGM output. (The complete output image, with gene tracks and variants table is shown in Supplementary **Figure S2**.) **C**: OGM identified two monomodular alleles in Participant #1. In the top haplotype, each of the three module-specific labels of the reference mapped to a single label in the sample, suggesting a monomodular haplotype (with HERV-K) carrying either the gene or the pseudogene. A 32,685 bp deletion was called (See Supplementary **Figure S3**). In the bottom haplotype, in the complete absence of HERV-K, the software called a ∼39 kbp deletion and aligned the single *CYP21* label to the active gene, suggesting a deletion of the pseudogene. Black arrows were added to these Access software screenshots to locate the next label upstream in the RCCX module, located in *STK19*. **D**: Only mono- and bimodular haplotypes could be identified in the OGM software outputs. Bimodular included reference (found in heterozygous state in Proband#7 and Father#2) and reference minus HERV-K in the second module (termed Hap1; heterozygous in Probands #3 and #7). The most common was a monomodular haplotype with a single, unidentified copy of *CYP21* (Hap 2), found in heterozygous state in Participant #1, Proband #3, Father #2, and in homozygous state in Proband #2 and his mother. Another shorter monomodular haplotype was seen in heterozygous state in Participant #1. In all, the *TNXB*-specific label was preserved. **E**: When aligned to hg38, in this proband (#2) homozygous for the monomodular Hap2, the single label for *CYP21* mapped to both *CYP21A2* and pseudogene (Supplementary **Figure S4**), as in other samples. But on hg19, it mapped unambiguously to the pseudogene (orange triangle and arrows), leaving the reference *CYP21A2* label (green triangle and arrow) unmapped. This suggested a complete deletion of the active gene. (Triangles were added only to the bottom allele; the top allele assembly was identical).

In **Proband #3**, with a clinically reported recurrent Int2G SNV and a “large gene deletion”, the OGM analytical pipeline identified two different haplotypes of the RCCX region (**Figure 3A**). One was bimodular, with deletion of the HERV-K in the second module. On the other haplotype a 32.6 kb deletion was called, with the centromeric *TNXB*-specific marker preserved. However, the remaining 3 labels mapped to both modules of the reference and the gene could not be distinguished from the pseudogene. This can be interpreted as a monomodular haplotype, but whether the gene or the pseudogene is missing could not be determined.

In **Proband #7** clinical testing had reported a heterozygous full deletion of *CYP21A2*, a hemizygous Int2G pathogenic variant in the remaining active gene, and three copies of the pseudogene detected with MLPA. While the method cannot resolve the cis/trans configuration, it suggests the presence of a trimodular haplotype carrying the SNV and a monomodular deletion haplotype.

Instead, OGM output showed 2 bimodular haplotypes with one *CYP21A1P* and one *CYP21A2* each (**Figure 3B** and Supplementary **Figure S2**). One haplotype harbored the HERV-K transposon in the first but not the second module. In the other haplotype the transposon was present in both modules, matching the reference. In both, the label located toward the end of the active *CYP21A2* gene was present and the clinically detected full gene deletion was not visualized.

In **Participant #1** OGM clearly identified two different haplotypes, both monomodular with only one carrying HERV-K. Each had only one label for *CYP21* but alignment to the reference behaved differently. In the haplotype with no HERK-V (bottom track in **Figure 3C**), the algorithm mapped the *CYP21* label to the active gene, and a 39-kb deletion was called (see Supplementary **Figure S3**). In the haplotype with HERV-K, the label mapped ambiguously to both the gene and the pseudogene (and the next label was upstream in the *STK* gene, black arrow). A 32-kb deletion was called, which could be a fusion allele or a complete gene deletion. Clinical testing identified only one pseudogene and one (wild type) gene in this unaffected sibling. So the number of modules was identified correctly by OGM and Hap1 may be the deletion allele (HERV-K, 1 pseudogene, no gene), and Hap2 may be a monomodular allele with only the intact gene.

Expected alleles in **Proband #2** include one Int2G variant and one large gene deletion. Information about the number of pseudogenes has been lost. Here the two haplotypes were indistinguishable (Supplementary **Figure S4**) and appeared identical to Haplotype 2 in Proband #3 (**Figure 3A**) or Hap1 in Participant #1 (Supplementary **Figure S3**, top). This appears to be a monomodular allele, with each label of the RCCX module mapped by the Access software to two positions on the reference assembly. A 32,581 kb deletion was called on each. Whether the pseudogene or the active *CYP21A2* is missing could not be determined.

The unaffected mother showed an identical pattern, with a 32,519 bp deletion called. The father appeared heterozygous with one haplotype similar to the proband’s and mother’s, and the other that aligned directly to the reference (bimodular with HERV-K in each copy of the *C4* gene).

In all we were able to distinguish four haplotypes, two monomodular and two bimodular, using OGM on these 7 samples (**Figure 3D**). The bimodular ones either aligned exactly to the reference (Proband #7, Father #2) or were missing HERV-K-in the second module (Hap1; found in Probands #3 and #7). Hap2, the most common haplotype (half of the 14) in this cohort was monomodular with a single label in *CYP21*, which did not allow distinction between gene and pseudogene. Another shorter monomodular haplotype was found in Participant #1.

Calls were different with some older assemblies, aligned to hg19 vs. hg38 references. For example, in Participant #1, on hg38, both HERV-K labels of the reference mapped to the single one in the sample. On hg19, only one did, and the other was left unmapped (compare Hap1 in the two panels of **Figure S3**), and the 32.5 kb deletion was not called (red arrows).

Similarly, the two reference labels shared by *TNXA* and *TNXB* co-mapped to the single one in Proband #3 on hg38 but not hg19 (pink arrow in Supplementary **Figure S5**). This did not impact SV detection as the same two deletions (6.5 kb and 32.5 kb) were called.

Interestingly alignment to hg19 yielded a more accurate call in one of the samples. In Proband #2 who appeared homozygous for the monomodular Hap2, on hg19, the single *CYP21* label mapped unambiguously to *CYP21A1P*, showing the *CYP21A2* label as missing (**Figure 3E**). This suggests a complete deletion of the gene, in accord with the clinical report.

### *Paraphase* phased deletions, fusions, and SNVs in the HiFi LRS dataset

HiFi sequence data sets were generated on a Revio instrument for six samples, three probands (including the above Proband#3) with clinically determined variants and a trio with no prior molecular testing. Analysis with PacBio’s WGS Variant Analysis pipeline accurately identified the Int2G variant in Probands #3, #4, and #6, as well as the nonsense Q319X variant in Proband #5. It phased the two SNVs in Proband #5 in compound heterozygote configuration and revealed a heterozygous Q319X variant in the proband and father of the undiagnosed Trio #4. It did not, however, call fusion or deletion alleles.

The output of the PacBio Paraphase tool, designed to disambiguate the alleles of genes of clinical relevance located in segmental duplications (X. Chen 2023, 2025), was then examined. The tool called the SNVs, located them in individual RCCX modules, ordered the modules in each haplotype, and identified and phased fusion and whole-gene deletion alleles. (Paraphase alignments of RCCX reads are visualized in IGV in **Figure 4**, with unambiguously mapped reads in lime or lilac for each haplotype and, in grey, reads who could align to more than one haplotype). Results were concordant with available clinical reports and, in addition, phased and calculated the number of modules for each haplotype, which may illuminate phenotype/genotype correlations but is not currently determined by clinical tests.

**Figure 4.**
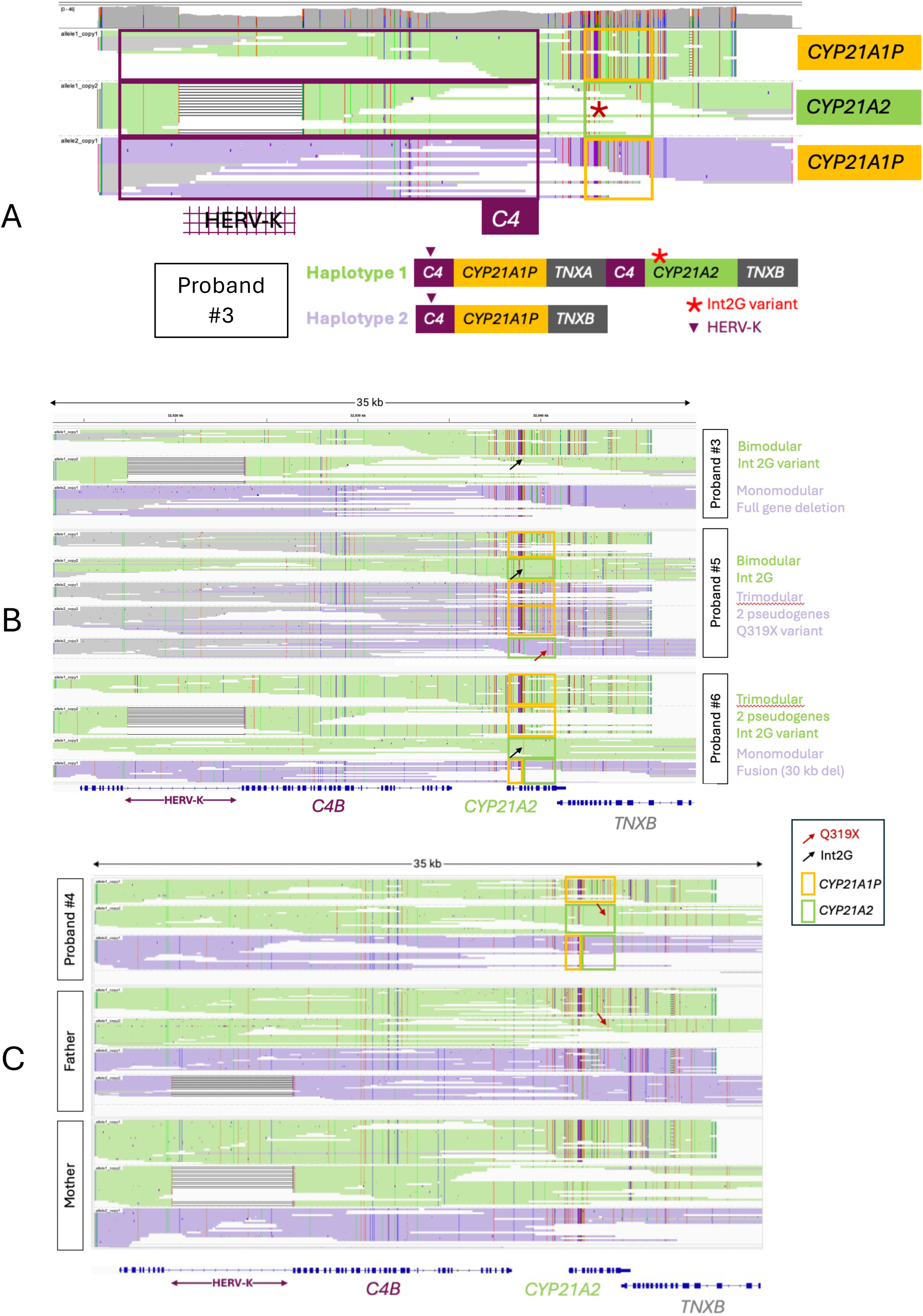
Paraphase resolved the haplotypes and pathogenic alleles of the RCCX module in six samples sequenced with HiFi LRS. **A:** Paraphase output of HiFi LRS dataset for **Proband #3** showed a bimodular haplotype (lime, see Haplotype 1 in Figure 3A for the same proband) with HERV-K in the first (top), but not second RCCX module, and a monomodular Haplotype 2 (lilac) with an intact pseudogene fused with the *TNXB* gene, *i.e.* a complete deletion of the active gene. The Int2G variant (asterisk) is in the second module of Hap1, in the active *CYP21A2* gene, in trans with the gene deletion allele. The mini-bams restricted to the region of interest are visualized in IGV. HiFi reads are realigned to the RCCX copy on GRCh38 that encodes *CYP21A2* and grouped into haplotypes. Reads in lilac or lime are consistent with a single haplotype. Reads in gray are consistent with more than one possible haplotype, *i.e.* when two or more haplotypes are identical over a region. Longer modules represent the last (centromeric) RCCX module, as they carry the long *TNXB* while shorter modules carry the truncated *TNXA* pseudogene instead. *C4*, *CYP21A1P* and *CYP21A2* are shown here framed in purple, orange, and green respectively. The multiple variants in the pseudogene clearly distinguish it from the active gene. The cartoon shown below the IGV output illustrates the *C4*, *CYP21* and *TNX* paralogs in the RCCX modules in this proband (with the same color conventions as in Figures 1 & 3). [The cartoons for the other samples are shown in **Table 1**]. **B**: Identification of biallelic pathogenic variants in Paraphase-resolved haplotypes in three affected individuals. A black arrow indicates the Int2G variant in Proband #3 (top panel) and others on the IGV visualization. RefSeq genes are shown at the bottom. The tracks from Proband #3 are shown again, without annotation rectangles for reference. In Proband #4 (middle panel), 5 modules were identified, in a bimodular haplotype (lime reads) carrying an Int2G variant in *CYP21A2*, and a trimodular haplotype (lilac reads), with duplicated pseudogene and a Q319X variant in the active gene (red arrow). In Proband #6 (bottom panel), a trimodular (lime) haplotype with HERV-K in the first and third modules carried an Int2G pathogenic variant in the *CYP21A2* gene in the third module. The other (lilac) haplotype was monomodular, with fusion of the 5’ end of *CYP21A1P* and the 3’ end of *CYP21A2* resulting in an inactive hybrid, characteristic of the clinically identified 30-kb deletion. **C**: Paraphase-resolved haplotypes in the previously undiagnosed Trio #4, showing biallelic pathogenic variants in the proband but not her unaffected parents. The (lime) bimodular allele carrying a Q319X variant was inherited from the father; the (lilac) monomodular allele inherited from the mother was a fusion of *CYP21A1P* to *CYP21A2* gene (as in Proband #6), likely representing the “30-kb deletion”.

In **Proband #3**, similarly to OGM, Paraphase identified one monomodular and one bimodular haplotype (**Figure 4A**). In addition, Paraphase was able to resolve the pathogenic alleles. In the bimodular haplotype, the first RCCX module (top lime-colored reads) contained the *CYP21A1P* pseudogene and HERV-K. In the second module, HERV-K was deleted and the *CYP21A2* gene carried the Int2G variant. The monomodular allele (shown in lilac) resulted from fusion of the *CYP21A1P* pseudogene to *TNXB*, with a full *CYP21A2* gene deletion.

**Proband #5** had undergone a limited clinical test that detected two pathogenic SNVs (Q319X and Int2G) but could not determine the number of pseudogenes or phase. HiFi LRS confirmed the two SNVs and phased them on a trimodular (with pseudogene duplication) and a bimodular haplotype respectively. Thus, Paraphase was able to disambiguate 5 modules, despite the high sequence similarity in some regions where some reads could not map unambiguously to a specific module (see grey reads, especially in the *C4* gene in **Figure 4B**, middle panel).

In **Proband #6** a trimodular haplotype with two pseudogenes and an active gene carrying the Int2 variant was detected (lime reads in **Figure 4B**, bottom panel). The other haplotype (lilac reads) was a fusion resulting in a *CYP21A1P*/*CYP21A2* inactive hybrid, which represents the so-called 30-kb deletion, as reported in clinical testing. We noted that, in the trimodular haplotype, the first pseudogene is lacking three variants for a “typical” *CYP21A1P* (V282L, Q319X, and R357W) but this was seen frequently in the population (It was 34.7%, 19.2% and 52.7% of *CYP21A1P* alleles lack V282L, Q319X and R357W, respectively when Paraphase was run on 259 individuals of five ancestral populations). These polymorphic sites in *CYP21A1P* could be a result of reverse gene conversion (fragment of *CYP21A2* moved to *CYP21A1P*) but, in any case, the remaining variants result in a functionally inactive copy.

In the undiagnosed **Trio #4** (**Figure 4C**), the proband inherited a (lime-colored) bimodular allele carrying the Q319X pathogenic variant from his father and a (lilac-colored) monomodular allele with a fusion *CYP21A1P/CYP21A2* hybrid from his mother. The other parental haplotypes were identical, bimodular, where the first module carried the pseudogene and HERV-K, and the second a wild-type *CYP21A2* and no HERV-K.

### Parakit is a new tool to resolve the RCCX region on Nanopore LRS datasets

The same 6 samples (3 clinically diagnosed singletons, 1 undiagnosed trio) were also subjected to nanopore-based LRS. In addition, two samples of Trio #2 failed extraction and only the mother could be processed. Assembly and variant calling using the Napu pipeline with default parameters could not originally identify or phase all pathogenic alleles. The reads were too short to span multiple RCCX modules, which confused assembly and phasing when aligned to the reference genome. More specifically, the original Napu pipeline was not able to call the deletion/fusion confidently and was only able to phase the pathogenic SNVs in two out of four probands (not shown).

To better analyze this challenging segmentally duplicated region, we developed a new tool, Parakit, which first builds a local pangenome representation, where the paralogous modules are collapsed (using reference modules from GRCh38 and alleles from the Human Pangenome Reference Consortium). Local long reads in each patient dataset were then extracted and realigned to this local pangenome. From these alignments, Parakit was able to identify and visualize pathogenic alleles in compound heterozygous configurations for all four probands. The pathogenic SNVs were found in multiple reads by the *variant-finder* command, which extracts variants from the ClinVar database, and in the haplotypes predicted by the “**haplotype-finder**” command. Deletions/fusions were found by those commands and further supported by the read coverage and allelic ratio changes displayed with the “**allele-support**” command. **Figure 5** shows the visualization of all these tools’ outputs for Proband #6.

**Figure 5:**
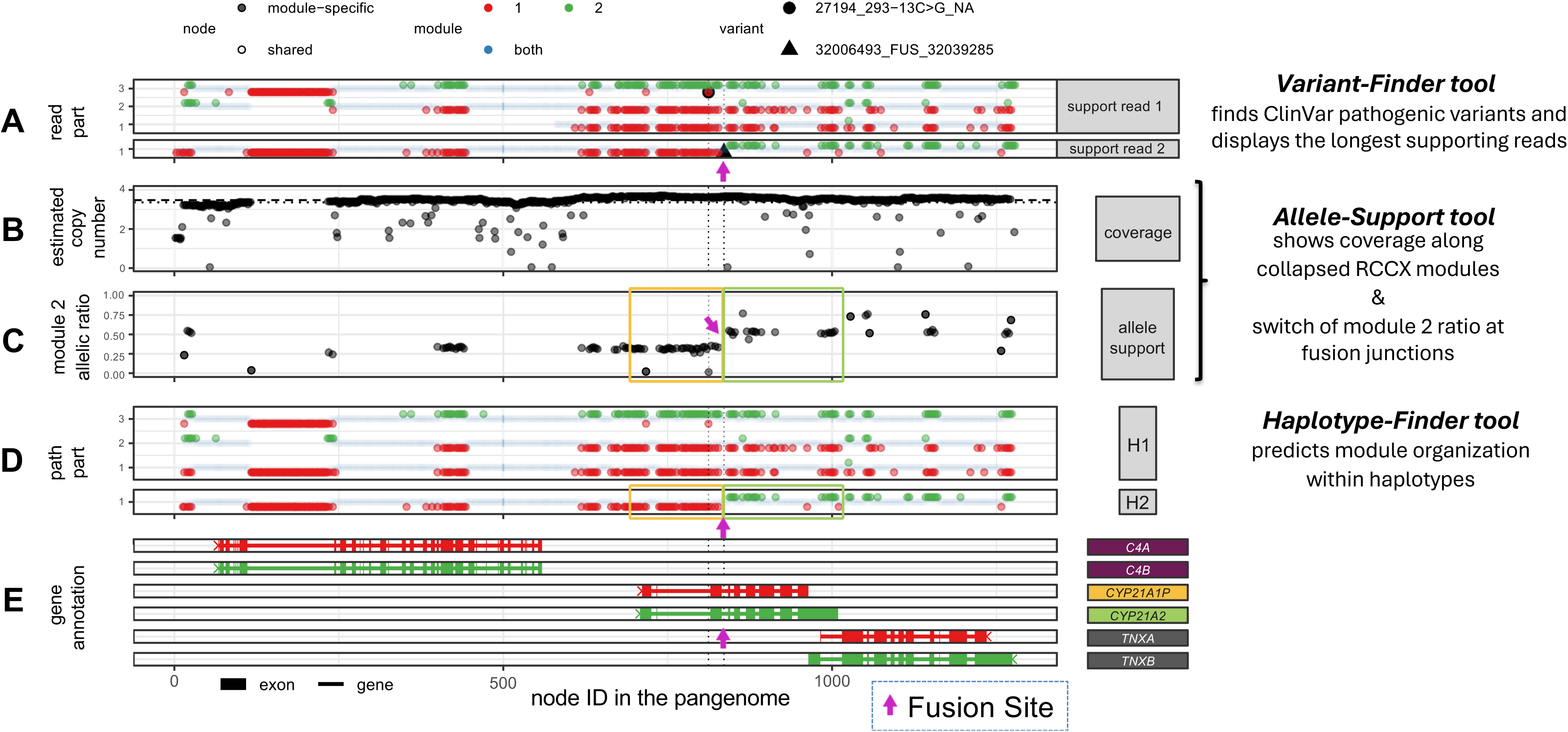
Summary visualization of the different analyses from Parakit for Proband #6. All panels share the x-axis representing the node position in the pangenome, which can be interpreted as a position in the consensus of the paralogous modules. A pink arrow indicates the fusion site in all panels. **A**: Output of the **“Variant-Finder”** command, which identifies ClinVar pathogenic variants and novel fusions, and displays the longest reads supporting them (here one read per allele). The y-axis separates the reads in parts when they traverse multiple RCCX modules and cycle through the pangenome. Informative nodes are colored in red and green, when they are markers of module 1 or 2, respectively. The Int2G variant and a fusion occurring in intron 3 are shown with a circle and triangle respectively. Vertical dotted lines from these events serve as alignment guides across all panels of Parakit’s output. Coverage (**B**) and the allelic support ratio of module 2 (**C**) are output by the **“Allele-Support”** command. In **B**, across most of the module, the aggregated read coverage is between 3 and 4. The horizontal dotted line highlights the estimated coverage from the median coverage and from the coverage around the cycling nodes (see Supplementary Methods). In **C**, the ratio shifted from about 0.25 (suggesting the presence of one *CYP21A2* and three *CYP21A1P* modules) to 0.5 (same number of *CYP21A2* and *CYP21A1P* copies) after the breakpoint of the predicted fusionThis is consistent with a hybrid fusion 5’-*CYP21A1P/CYP21A2*-3’. **D: “Haplotype-Finder”** command output. Similarly to how reads are represented in **A**, predicted haplotypes traverse the pangenome through module-specific markers (red/green points). Haplotype H1 shows a trimodular haplotype with, from the telomeric end, a module with HERV-K and *CYP21A1P*, as second one with *C4* without HERV-K and another copy of *CYP21A1P*, and a third module carrying *CYP21A2* with the Int2G pathogenic variant. The second predicted haplotype was a fusion, occurring intron 3, visualized with change of node color from red to green. **E**: Position of the genes and pseudogenes of the RCCX module in this pangenome scale to help position the variants within the modules. The thicker rectangles represent exons, and an arrowhead the start site.

In **Proband #6**, Parakit detected the Int2G pathogenic variant on a trimodular haplotype and a fusion in intron 3 (*i.e.* deletion of the first two exons of *CYP21A2*) leading to an inactive 5’-*CYP21A1P/CYP21A2*-3’ hybrid, characteristic of the 30-kb deletion. This was entirely concordant with the clinical test and with the Paraphase findings on the HiFi dataset. The top panels of the Parakit output show the ClinVar pathogenic variants identified by the ***variant-finder*** command, and the longest reads supporting them. The reads are split in multiple “parts” (y-axis) if they traverse the pangenome multiple times. Each dot represents a node in the pangenome traversed by the reads and is colored in red (or green) if a specific marker of module 1 (or 2). Here, the supporting reads were mutually exclusive, suggesting that the two variants are in compound heterozygous configuration (**Figure 5A**). We also notice that the longest read supporting the pathogenic SNV traverse three modules (3 parts), suggesting the presence of a trimodular allele.

The ***allele-support*** command shows the allelic ratio of module 2, with an expectation of around 0.5 for reference bimodular alleles. The ratio would be expected to be ∼ 1/3 and 2/3 for tri-modular alleles with two *CYP21A1P* or two *CYP21A2* respectively. In this sample, the ratio shifted from about 1/4 (suggesting the presence of 1 *CYP21A2* and 3 *CYP21A1P* modules) to 1/2 after the breakpoint of the predicted fusion occurring in intron 3 of *CYP21A2* (see pink arrow in **Figure 5C**). As the coverage remained unchanged at the fusion point, this is also consistent with a 5’-*CYP21A1P/CYP21A2*-3’ hybrid.

The ***haplotype-finder*** command outputs represent the pairs of haplotypes predicted from the read alignment through the pangenome. The same representation is used as for reads to show the path taken by each haplotype in the pangenome. Here 5 modules, a fusion and Int2G SNV were predicted, the latter within the last module of a trimodular haplotype creating a pathogenic allele (**Figure 5D**).

Similar evidence was observed for **Trio #4** with a fusion breakpoint at the same location as in Proband #6 and Q319X as pathogenic SNV in trans (**Figure 6A**). The parental haplotypes (Supplementary **Figure S6)** were consistent with the two variants predicted in the proband, with the mother carrying the fusion allele and the father the pathogenic SNV. The other, non-pathogenic, haplotypes in the parents were bimodular haplotypes with an absent HERVK in the second module, consistent with HiFi results (**Figure 4C**).

**Figure 6:**
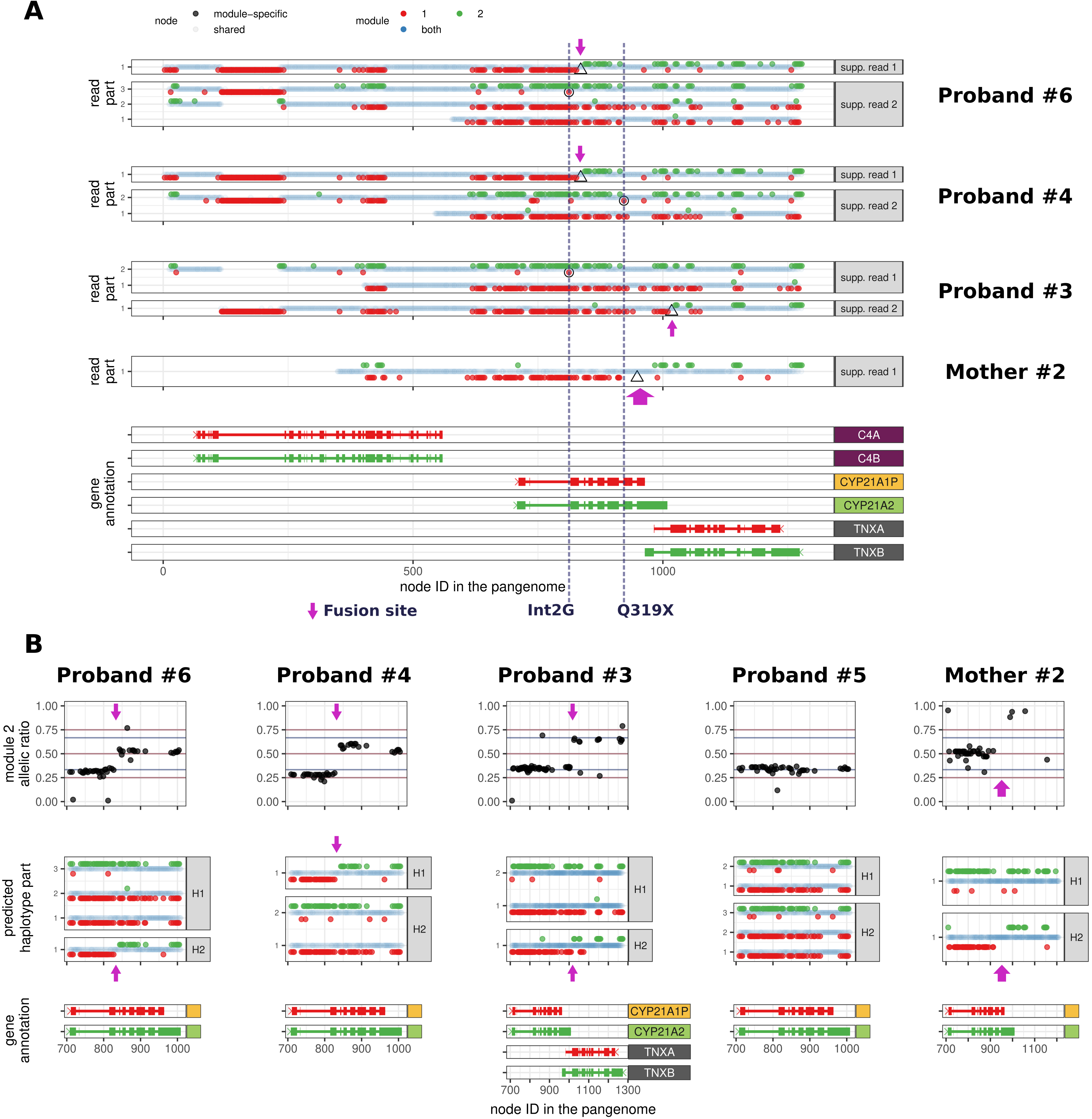
Visualization of fusions across samples with Parakit. **A:** Visualization of fusions with the *Variant-Finder* output. The longest reads supporting each pathogenic allele are shown as they traverse nodes along the pangenome (x-axis) for one sample in each family. Nodes colored in red or green are informative markers specific to module 1 and 2, respectively. ClinVar SNVs called by Parakit are highlighted with circles and the vertical dotted line. Fusions are highlighted with triangles and further annotated here with pink arrows. The wider arrow for Mother #2 illustrates the uncertainty of location of the fusion in this lower-quality dataset. The gene annotation track at the bottom allows to easily map the fusions in the various samples in relation with the *CYP21A2* and *TNXB* genes. (Proband #6 panels were repeated here and below from Fig. 5 to allow comparison with the other samples.) **B:** Visualization of fusions in the *Allele-Support* and *Haplotype-Finder* outputs. The *allele-support* track for each sample (top panels) shows a switch at the sites of predicted fusions (pink arrows). In Proband #3 and #4 module 2 allelic ratio switched from 1/3 to 2/3 after a fusion site occurring beyond the 3’ end and in Intron 3 of *CYP21A2* respectively. This suggests a fusion in the distal module. The switch from “red” (“*CYP21A1P*” module) to “green” (“*CYP21A2*” module) nodes occurred at the same positions in the visualization of the *haplotype-finder* output. For both, the fusion was predicted to be on a mono-modular haplotype, while the SNV was on a bimodular haplotype with one pseudogene and one gene. In contrast, in Proband #5, which carries two SNVs and no fusion, module 2 ratio remained similar throughout the region and nodes are the same color throughout each module. The ratio was approximately 0.4, which is compatible with the prediction of 5 RCCX modules in this sample, as shown in the output of the *haplotype-finder* tool (path part panel). A trimodular haplotype with two pseudogenes and a bimodular haplotype were predicted, as with the HiFi dataset. For Mother #2, the *haplotype-finder* output predicted two monomodular haplotypes, one carrying *CYP21A1P*, the other *CYP21A2.* Accordingly, module 2 ratio was at 0.5 until the predicted, increasing to 1 beyond. Here the precise location of the fusion could not be determined, and the width of the pink arrow reflects the location uncertainty.

For **Proband #3**, the *variant-finder* command found the pathogenic Int2G SNV and a fusion (**Figure 6A**), which was also supported by the *allele-support* panel with a clear switch from 1/3 to 2/3 module 2 ratio around the fusion breakpoint (**Figure 6B**, Supplementary **Figure S7**). Unlike in the previous two samples, here the breakpoint was at the end of the *CYP21A2* gene, *i.e*. in the *TNXB* gene, and would result in a complete deletion of the active gene and fusion of *CYP21A1P* to *TNXB*. The *haplotype-finder* tool also supported the presence of both variants, as well as their compound heterozygous configuration.

The two pathogenic SNVs in **Proband #5** were found by the *variant-finder* and the haplotype prediction commands (**Figures 6A, 6B**). Here, even though none of the reads were long enough to traverse multiple modules, the *haplotype-finder* command was able to identify 5 modules (compare ‘read parts’ and ‘path parts’ tracks in Supplementary **Figure S8**). The copy number was estimated at 4.5 based on the read coverage around the cycling nodes and the module 2 ratio was below 0.5 across most of the module, both of which further support the presence of a trimodular haplotype with two pseudogenes as predicted by the *haplotype-finder* output. Each haplotype was predicted to carry only one copy of the active gene with the two SNVs in trans. The Int2G SNV was predicted on the bimodular haplotype, while the Q319X was placed on the trimodular haplotype. These biallelic pathogenic variants establish a firm diagnosis that the limited clinical testing this patient had undergone could not. In addition, Parakit *haplotype-finder* output suggests a *C4* gene conversion in the second module of the trimodular haplotype, which appears to carry *C4B* rather than *C4A* (**Fig. S8**).

For **Trio #2**, only the HMW DNAs previously extracted for OGM were available to use for LRS. Proband and father samples failed library preparation, and mother yielded the lowest coverage of the entire cohort (**Table S2**). It was still sufficient for Parakit to decipher the haplotypes. The *variant-finder* output identified a single pathogenic fusion, at a different location than in Probands #4 and #6 (**Figure 6A**), deleting most of the *CYP21A2* gene (Supplementary **Figure S9**). The outputs of the *allele-support* (Estimated copy number = 2; Allelic ratio of module 2 shift from 0.5 to 1 at the point of fusion) and of the *haplotype-finder* tools were consistent with the presence of two mono-modular haplotypes, one carrying only the active gene, and the other a fusion deleting most of the gene. Exact fusion location could not be further determined in this low-quality sample.

The Parakit visual output made it easy to compare the location of the breakpoints for the fusions (**Figure 6A**). Calculation of “Module2 ratio” (*i.e.* the fraction of modules containing *CYP21A2*) also showed a change of value at the points of fusion (**Figure 6B**) and total coverage helps assess the total number of modules.

## Discussion

DLE-1 labeling didn’t provide enough markers in the region for OGM to confidently distinguish CAH pathogenic recombination alleles. The Access software did not provide a way to calculate the number of modules, and we were not able to reliably interpret this segmentally duplicated region. Development of other labeling strategies or specific bioinformatic tools (like Paraphase and Parakit for LRS) may allow distinguishing the gene and pseudogene in the future. The presence or absence of the Herv-K retrovirus in the *C4* gene was reliably detected, which may be useful for other studies. The discrepancies observed between hg38 and hg19 in some, but not all, haplotypes suggest that the OGM dataset may be richer than the output suggests, and that it could be mined for more accurate calls in future versions of the software.

The two LRS technologies we tested, PacBio HiFi and ONT nanopore-based sequencing, were able to identify and phase the various complex alleles in the *CYP21A2* region and could replace the current cumbersome clinical molecular test for CAH. The usefulness of LRS to disambiguate pathogenic alleles in 21-hydroxylase deficiency was recently demonstrated using targeted capture of the *CYP21A2* region in Chinese (Zhang 2024) and Japanese (Adachi 2023) cohorts.

However, we could not determine the specificity of variant identification in those studies, and the approach was not reported to allow detection of *TNXB* deletions. Another report (Liu 2022) used LRS but still required the long-range PCRs to amplify the region. In contrast, a long-read whole-genome sequencing-based approach using the strategy we deployed here would not only provide more complete genotyping of the entire RCCX module, including information on the other genes in the module beyond *CYP21A2*, but will also allow detecting the rarer etiologies of CAH, associated with genes on other chromosomes, in a single test. Deployment of LRS in clinical settings is currently limited by cost and amount of DNA required. However, this is likely to change rapidly in the future. In November 2024, PacBio announced the new SPRQ chemistry, with reduced DNA input requirements, which should decrease price per sample.

*Paraphase* is available to genotype the RCCX region in PacBio HiFi datasets (X. Chen, 2023, 2025). Taking advantage of pangenome data, we developed the computational suite, *Parakit*, to call the various pathogenic alleles on Nanopore datasets. *Parakit* starts by building a local pangenome where the different paralogous modules of the Human Pangenome Reference Consortium are collapsed (Liao 2023). Local long reads are extracted and realigned to this pangenome. Four different approaches are then used to confidently call the various alleles: read coverage, the fraction of reads supporting variants between copies, the identification of putative junctions from long reads, and the reconstruction of full-length haplotypes. Parakit is provided with the local pangenome for the entire RCCX module. This could allow better genotyping of the region to clarify emerging genotype phenotype correlations for conditions linked to the other genes in the module, such as *C4* and schizophrenia or susceptibility to autoimmune disorders (see Proband #5, Fig. S8). Parakit can also be further adapted for any other segmental duplications, by creating a local pangenome for any region of interest.

Determination of the number of copies of the *CYP21A2* gene in cis is critical for an accurate diagnosis. For example, the p.Q319X variant has been shown to exist in the population, as the result of a founder effect, in one copy of the *CYP21A2* where the gene is duplicated (Kleinle 2009). This is a non-pathogenic allele, as the second copy of the gene is active. Hence, detecting only the SNVs without counting the modules can lead to false positives (Lekarev 2013). We did not have an example of this in our dataset, but Paraphase has been shown to detect this common haplotype as well (X. Chen 2025). Where the variant was found here (probands #4 and #5), it was in a trimodular haplotype with 2 copies of the pseudogene and only one of the active *CYP21A2* gene, resulting in a pathogenic allele consistent with phenotype.

Precise information about the number of modules on each allele may also inform further genotype-phenotype correlations in 21-hydroxylase deficiency. While specific variants are reported to be associated with the different forms of CAH (*e.g.* the 30-kb deletion and Int2G with the severe, salt-wasting form), genotype-phenotype correlations are not absolute (Speiser 1992; Krone 2000; Kim 2023). The largest (∼1,500 samples) study to date concluded that only 21 of the 45 genotypes examined yielded an absolute phenotypic correlation, undermining prediction of newborn phenotype from prenatal mutation testing (New 2013). However, it is worth noting genotyping was limited to 8 common variants (by Sanger sequencing of PCR-amplified fragments) and the “large deletion” (on Southern blot) in that study. Reexamination of these results with genotyping of the entire region may lead to better predictive value for each allele.

In patients with 21-hydroxylase deficiency CAH who also carry a deletion of *TNXB*, a chimeric *TNXA/TNXB* gene causes tenascin-X haploinsufficiency. The contiguous gene syndrome has been termed CAH-X, and associates symptoms of hypermobility-type Ehlers Danlos Syndrome with CAH (Merke 2013; Gao 2020; Marino 2021; Concolino & Falhammar 2022; Kim 2023). Such variants should be identified early to allow for the detection and management of muscular and cardiovascular complications linked to abnormal expression of TenascinX. Having this information included in the clinical test for CAH would alert the endocrinologists to convene an interdisciplinary team to assess other features of the syndrome. Another finding awaiting explanation is a possible sex difference observed in CAH-X syndrome: among 20 obligate heterozygotes for a severely defective *TNXB* allele, 9 of 14 females but no males had hypermobility (Zweers 2013). Examination of the PacBio dataset suggested that *TNXB* is intact in all the samples in our study, and none of the patients have reported symptoms to date.

In addition, the non-classic form of CAH is common in the population but mostly undiagnosed, or misdiagnosed with other causes of hyperandrogenism, such as polycystic ovary syndrome (Escobar-Morreale 2008; Speiser 2009; Papadakis 2019). However, accurate diagnosis is important for both management and family planning. For example, treatment with glucocorticoids, and/or birth control medications may be beneficial to improve fertility and manage signs of hyperandrogenism (Eyal 2017; Livadas 2019; Carrière 2023). A convenient, reliable clinical test supported by LRS should facilitate correct diagnosis.

To record the genotypic complexity of this region newly made accessible with LRS, the DSD-TRN has created a new standardized form for clinical data collection for CAH. It is available in REDCap or as a spreadsheet. It documents testing laboratory, test performed, pathogenic alleles, phase, number of pseudogenes/modules, in probands and family members, CAH phenotype, as well as genetic status of *TNXB*. It will be expanded to guide the clinical assessment of TenascinX-deficiency features to support deep phenotyping of CAH-X.

## Data Availability

The RCCX pangenome and pipeline to perform the described analysis are included in the Parakit repository in GitHub. HiFi Paraphase output and other data are available upon reasonable request to the authors.

https://github.com/jmonlong/parakit

## Supplementary Material

- Parakit detailed methods

- Supplementary Tables S1-S2

> Table S1: Participants’ clinical tests, phenotype, demographics (information withheld per medrxiv policy)
>
> Table S2: Metrics of the Hifi, Nanopore, and OGM datasets

- Supplementary Figures S1-S8

> S1 IGV visualization of OGM DLE1-labeled CTTAGG sequence
>
> S2 OGM output aligned to hg38 for Proband #7
>
> S3 OGM output aligned to hg19 and hg38 for Participant #1
>
> S4 OGM output aligned to hg38 for Trio #2
>
> S5 OGM output aligned to hg19 and hg38 for Proband #3
>
> S6 *Parakit* output for Trio #4 (nanopore-based LRS)
>
> S7 *Parakit* output for Proband #3 (nanopore-based LRS)
>
> S8 *Parakit* output for Proband #5 (nanopore-based LRS)
>
> S9 *Parakit* output for Mother #2 (nanopore-based LRS)

## Acknowledgements

We are very grateful to the families who entrust us with their samples for research purposes. Dr. Jonah Cool from the Chan Zuckerberg Initiative was instrumental in getting the project launched and funded. Cairbre Fanslow, Christine Lambert, and Primo Baybayan of Pacific Biosciences were responsible for HiFi library preparation and sequencing. We thank Dr. Surajit Bhattacharya, Children’s National Research Institute, for discussions of OGM data.

## GitHub repositories

○ **Parakit** https://github.com/jmonlong/parakit
○ **Phrank Python implementation pipeline for HiFi** https://github.com/PacificBiosciences/wdl-dockerfiles/blob/main/docker/wgs_tertiary/scripts/calculate_phrank.py
○ **PacBio WGS Variant Pipeline v1.1.0** https://github.com/PacificBiosciences/HiFi-human-WGS-WDL
○ **Paraphase** https://github.com/PacificBiosciences/paraphase

## Ethics declaration

Generative AI was not used to write this manuscript.

Informed consent was obtained by the referring clinical teams for families to provide deidentified clinical data and blood samples to the DSD-TRN. Ethical approval was granted by the following: Internal Review Board of the Ann & Robert H. Lurie Children’s Hospital of Chicago (Protocol #2015-536), the Colorado Multiple Institutional Review Board at the University of Colorado (Protocol #19-3084), Northwell Health Internal Review board (Protocol #15-001), and Institutional Review Boards of the University of Michigan Medical School (IRBMED, Protocol HUM00050916)

Use of the deidentified DSD-TRN biobank samples for genetic research was approved by the Institutional Review Board at Children’s National Hospital, Washington, D.C., USA under protocol P000010217.

## Authors contributions

Conceptualization ED, EV, SIB, BP

Data curation: HB, XC, ED, SN, BM, XC

Funding acquisition EV, KM, BP, SIB, ED

Investigation: HB, XC, BM, SN, WJR,

Methodology: JM, BP, KM, BM, XC, WJR

Project administration: ED, MA, ID

Resources: AR, BP, EV, PWS, CF, CEK, JH, LM, NN

Software JM, XC

Supervision KM, BP, AR, ED, PWS

Visualization ED, JM, XC

Writing-original draft ED

Writing-review & editing ED, JM, BP, ID, PWS, NN, XC, WJR

