## Supplementary methods, tables, & figures for "Long-read sequencing resolves the clinically relevant *CYP21A2* locus, supporting a new clinical test for Congenital Adrenal Hyperplasia"

### Supplementary Material

- Parakit detailed methods
- Supplementary Tables S1-S2

    Table S1: Participants' clinical tests, phenotype, demographics (information withheld per medrxiv policy)

    Table S2: Metrics of the Hifi, Nanopore, and OGM datasets

- Supplementary Figures S1-S8

    S1 IGV visualization of OGM DLE1-labeled CTTAGG sequence

    S2 OGM output aligned to hg38 for Proband #7

    S3 OGM output aligned to hg19 and hg38 for Participant #1

    S4 OGM output aligned to hg38 for Trio #2

    S5 OGM output aligned to hg19 and hg38 for Proband #3

    S6 *Parakit* output for Trio #4 (nanopore-based LRS)

    S7 *Parakit* output for Proband #3 (nanopore-based LRS)

    S8 *Parakit* output for Proband #5 (nanopore-based LRS)

    S9 *Parakit* output for Mother #2 (nanopore-based LRS)

#### Parakit detailed methods

Files and commands to reproduce the analysis shown in this study are documented in https://github.com/jmonlong/parakit/tree/main/paper. We built a pangenome for the RCCX region that can contain a module with the *CYP21A1P* pseudogene (module *P*), and a module with the *CYP21A2* gene (module *G*).

##### *Pangenome construction*

Parakit will work with the sequence of the region of interest and the module *P* and *G*. Hence, the first step is to specify the coordinates of both modules in GRCh38 (chr6:31980532-32013273 for module *P*, chr6:32013273-32046127 for module *G*) and the size of the flanking region to include (300 kbp). Parakit starts by creating a reference sequence without module *G*, to use as a backbone for the pangenome. It then creates a pangenome using Minigraph-Cactus, augmenting the backbone with sequences representing the module *G* extracted from GRCh38, and module *P* and *G* extracted from 65 high-quality assemblies from the HPRC. The HPRC assemblies were selected because both the CAT and Ensembl gene annotations consistently identified one module *P* and one module *G*. Of note, Parakit also includes the sequence of module *P* and *G* from GRCh38 (here the sequence corresponding to chr6:31980532-32046127) as an input sequence for Minigraph-Cactus, to make sure there is an edge from the end of the collapsed module to its beginning. This cycling edge is necessary to allow a read or haplotype to traverse multiple modules. The full GRCh38 reference path is added to the pangenome using the *augment* subcommands of the vg toolkit.

After building the pangenome, Parakit annotates the nodes based on the number of module *P* and *G* traversing them. A node is marked as specific to module *G* if it is traversed by at least 3 times more module *G* than module *P*, and vice versa.

The command used to construct the RCCX pangenome with Parakit was:

```
parakit construct -j rccx.grch38_hprc.mc.config.json
```

The JSON configuration file contains information about the location of the GRCh38 reference FASTA file, coordinates of each module, size of the flanking regions, location of the HPRC assemblies.

##### *Alignment of long-reads to the pangenome*

Parakit first extracts reads in the region of interest from an indexed BAM. This assumes that the reads were first aligned to the reference genome, for example using minimap2. Although the read alignment around the RCCX module might not be accurate, reads of interest should still be mapped to the region. Hence, we should retrieve all informative reads by extracting those that were originally mapped to this region of the linear reference.

Once retrieved, the reads are re-mapped to the pangenome using GraphAligner. Internally, Parakit uses GraphAligner v1.0.17 with variation graph mode (-x vg) and 100 bp for the alignment bandwidth (-b 100).

To extract and map long reads to the pangenome with Parakit, we ran:

```
parakit map -j rccx.grch38_hprc.mc.config.json -b SAMP.wgs.bam -o
SAMP.rccx.grch38_hprc.mc.gaf.gz
```

The output is a GAF file representing the alignment of each read through the pangenome. Because the paralogous regions are collapsed, each read maps confidently to only one position in the pangenome. A long read can often traverse the collapsed region of the pangenome multiple times when it spans multiple modules.

##### *Read-based variant calling*

Aligned reads that traverse the pangenome through module-specific nodes (as defined above) can be used for inference. In Parakit, the call subcommand searches for evidence of gene fusion or gene conversion in each read separately. Specifically, a sliding window approach looks for positions - within the RCCX module - where a read switches from aligning to *P*-specific nodes to aligning to *G*-specific nodes. We used sliding windows of 20 informative markers on each side of a potential variant site. For fusions, we select sites where at least 80% of markers in the upstream window are specific to module *P*, and 80% of markers in the downstream window are specific to the module *G*. For small gene conversion event, we look for module *P* nodes, specifically those corresponding to known ClinVar variants, surrounded by module *G* nodes. Here, Parakit selects candidates if there are more than three times more module *G* nodes than module *P* in the windows upstream and downstream. For both fusions and gene conversion variants, Parakit reports sites with at least 3 supporting reads.

To call variants with Parakit, we ran:

```
parakit call -r SAMP.rccx.grch38_hprc.mc.gaf.gz -j
rccx.grch38_hprc.mc.config.json -o SAMP.rccx.grch38_hprc.mc.calls.tsv
```

The ClinVar database was downloaded on Sept 3, 2024 from https://ftp.ncbi.nlm.nih.gov/pub/clinvar/tab_delimited/variant_summary.txt.gz.

##### *Coverage and allele ratio along the pangenome*

Parakit uses the read alignment through the pangenome to compute estimates of coverage and a ratio of *G* alleles along the RCCX region.

Read coverage helps estimate the total number of module copies. We count the number of reads covering each of the non-module-specific nodes (light blue points in the figures). These nodes are not specific to a module so they should be traversed by all reads, not matter the allele, and provide an estimate of total copy number. This coverage can be plotted along the pangenome (see Visualization of the results below). Parakit also computes two global copy number estimates. The first is simply the median read coverage across the non-specific nodes, as mentioned above, normalized by the coverage in flanking regions. We note that this approach can underestimate the copy number when there is mapping bias in some parts of the region (typically lower coverage in the middle of the RCCX module). The second copy number estimate focuses on the reads entering (or leaving) the collapsed region, and the reads cycling back from the end of the module back to the beginning. Indeed, each cycle supports the presence of an additional copy. Parakit estimates the copy number as 2*(Rf+Rc)/Rf where Rf is the number of reads entering (or exiting) from the flanks and Rc is the number of cycles supported by reads.

Parakit also helps look for changes in the allelic balance at informative sites, i.e. the proportion of reads traversing nodes specific to module *G*. Changes in this proportion is expected around breakpoints of a fusion allele or at the boundaries of a large gene-converted region. Each module-specific node is assigned an anchor node as the first non-specific reference node upstream. For each anchor node, we compute the coverage of *G*-specific nodes divided by the coverage of nodes specific to either *P* or *G*. In the presence of two bimodular haplotypes, the ratio should be centered around 0.5. An individual carrying a fusion haplotype and a bimodular haplotype (one module *P* and one module *G*) will show consistent allele *G* ratio around 1/3 up to the fusion breakpoint, and around 2/3 afterward. This evidence is orthogonal to the read-level evidence extracted by the variant calling or haplotype reconstruction approaches.

Both of those analyses are performed by the visualization command (see below) using the read alignment information.

##### *Haplotype reconstruction*

Parakit also infers the most likely pair of haplotypes (or diplotype) by finding the pair of paths through the pangenome graph that are the most consistent with the aligned reads. Haplotype reconstruction is performed in two steps: 1) haplotype candidates are enumerated, 2) the best pair of haplotype candidates is identified.

Haplotype candidates are first enumerated. The goal is to produce an extensive list of potential haplotypes guided by the read alignments. This is also done in two steps: module candidates are constructed and then stitched together into haplotypes. Read alignments are first split at the boundaries of the collapsed region of the pangenome. For example, a read spanning the flanking region and two modules will be split in three subreads: the upstream flank, the first module, and the second module traversed. All the subreads are then clustered using an iterative approach. All subreads start in the same cluster and a consensus path is produced. Parakit then looks for positions in this consensus where at least three reads disagree. If such variant markers are found, they are used to split the cluster in two. To do so, a network is built where reads are connected by edges that are weighted by the number of variant markers they have in common. The reads are split in two sets using the Kernighan–Lin algorithm, implemented in the network Python package. This process is iterated until no variant markers are found in the clusters. The consensus path for all clusters (and all iterations) are saved and combined in the final step into candidate haplotypes. Here, Parakit uses information about which subreads are assigned to a cluster and how the subreads were originally split to suggest adjacency between the clusters. Haplotypes are created by starting at the upstream flank and adding clusters if at least one read supports this adjacency, until it reaches the downstream flank or the maximum allowed number of modules is exceeded (currently set to 5).

The enumeration of the haplotype candidates described above is permissive on purpose to maximize the chances of containing the correct ones. In the second step of the haplotype reconstruction, Parakit looks for the pair of haplotypes that best match the read alignments, both in terms of read identity and read coverage. Read identity is measured as the average identity of the best alignment on the pair of haplotype candidates. This identity is computed in pangenome space, i.e. representing the proportion of nodes matching when aligning the reads and haplotype. Hence, this pangenome alignment identity gives more weight to variants than sequence alignment because (potentially long) stretches of common sequences are merged into single nodes. Read coverage uniformity is measured as the average deviation of the read coverage in each node compared to the expected coverage. The expected read coverage is computed based on the number of times the node is present in the tested pair of haplotypes. Both the identity and read coverage metrics are normalized by the maximum values across all evaluated pairs, summed and used to rank the pairs of haplotypes. The haplotype pair with the highest normalized score is selected as the most likely diplotype.

In summary, reads are represented by their traversal of the pangenome, naturally putting more weight on known variants, and used to predict potential haplotypes. The pair of haplotypes that results in the highest read identity and most uniform read coverage is then reported by Parakit as the most likely diplotype. The command to reconstruct haplotypes was:

```
parakit diplotype -j rccx.grch38_hprc.mc.config.json -r
SAMP.rccx.grch38_hprc.mc.gaf.gz -o SAMP.rccx.grch38_hprc.mc
```

##### *Visualization of the results*

Parakit uses R and the ggplot2 package to display the different layers of evidence described above on the collapsed pangenome. The different panels are horizontally aligned so that sites of variation (e.g. fusion, SNVs) can be compared easily between analyses. For example, to help check that the fusion predicted by reads is concordant with the switch in module G ratio and the predicted fusion in the reconstructed diplotype. It includes a gene annotation track with the exons and introns for the genes in the region. The command used to generate a PDF file with the graph was:

```
parakit viz -j rccx.grch38_hprc.mc.config.json -r SAMP.rccx.grch38_hprc.mc.gaf.gz -c
SAMP.rccx.grch38_hprc.mc.calls.tsv -d SAMP.rccx.grch38_hprc.mc.paths-stats.tsv -p
SAMP.rccx.grch38_hprc.mc.paths-info.tsv -o SAMP.rccx.grch38_hprc.mc.pdf
```

#### Table S2: Metrics of the Hifi, Nanopore, and OGM datasets

##### HiFi PacBio dataset

| Participant | Affected | Sex | relationship | num_reads | read_length_mean | read_length_median | quality_mean | quality_median | mapped_fraction | depth_mean | SNV | indel | Ts_Tv | het_hom |
|---|---|---|---|---|---|---|---|---|---|---|---|---|---|---|
| Proband #3 | TRUE | FEMALE | proband | 5,637,074 | 18,931.51 | 18,365 | 32.49 | 32.0 | 0.99987 | 34.23 | 4,432,366 | 983,148 | 1.80 | 1.45 |
| Proband #4 | TRUE | FEMALE | proband | 6,676,888 | 15,736.51 | 15,076 | 33.38 | 33.0 | 0.99989 | 33.72 | 4,448,273 | 982,733 | 1.80 | 1.46 |
| Father #4 | FALSE | MALE | father | 7,233,654 | 17,136.52 | 16,594 | 32.00 | 32.0 | 0.99990 | 39.75 | 4,441,191 | 982,772 | 1.80 | 1.40 |
| Mother #4 | FALSE | FEMALE | mother | 6,391,824 | 17,889.48 | 17,373 | 31.88 | 31.0 | 0.99988 | 36.69 | 4,461,732 | 986,983 | 1.80 | 1.48 |
| Proband #5 | TRUE | FEMALE | proband | 5,367,785 | 18,231.55 | 17,793 | 31.63 | 31.0 | 0.99986 | 31.39 | 4,503,677 | 993,292 | 1.81 | 1.45 |
| Proband #6 | TRUE | FEMALE | proband | 5,995,270 | 17,272.18 | 16,705 | 31.92 | 32.0 | 0.99988 | 33.22 | 4,467,136 | 983,035 | 1.80 | 1.46 |
| | | | | **6,217,083** | **17,532.96** | **16,984** | **32.22** | | | **34.83** | | | | |

| Participant | INS | DUP | DEL | INV | BND | CN_DEL | CN_DEL_sum | CN_DUP | CN_DUP_sum | phased_basepairs | block_ng50 | cpg_sites_combined | cpg_sites_hap1 | cpg_sites_hap2 |
|---|---|---|---|---|---|---|---|---|---|---|---|---|---|---|
| Proband #3 | 13,187 | 510 | 9,466 | 105 | 78 | 25 | 7,170,000 | 13 | 3,482,000 | 2,560,949,767 | 684,431 | 29,017,605 | 25,135,364 | 25,112,023 |
| Proband #4 | 12,992 | 515 | 9,456 | 94 | 94 | 29 | 7,608,000 | 18 | 6,306,000 | 2,480,180,589 | 462,491 | 29,022,890 | 24,180,576 | 24,131,521 |
| Father #4 | 13,006 | 536 | 9,439 | 92 | 94 | 28 | 7,270,000 | 18 | 5,584,000 | 2,413,791,966 | 523,736 | 29,201,959 | 25,301,139 | 25,262,982 |
| Mother #4 | 13,119 | 508 | 9,390 | 99 | 84 | 37 | 9,332,000 | 13 | 4,660,000 | 2,565,799,525 | 616,835 | 29,033,986 | 25,657,654 | 25,634,394 |
| Proband #5 | 13,230 | 534 | 9,660 | 82 | 78 | 27 | 9,632,000 | 13 | 5,640,000 | 2,524,162,920 | 654,227 | 28,835,709 | 20,704,538 | 20,720,913 |
| Proband #6 | 13,147 | 491 | 9,568 | 94 | 108 | 22 | 5,312,000 | 19 | 6,558,000 | 2,517,520,381 | 538,447 | 28,912,326 | 22,197,145 | 22,143,735 |

| Cohort-wide statistics (n=98) | |
|---|---|
| number reads mean ± SD | 5,775,181±1,164,253 |
| read length mean± SD | 18,185 ± 2,610 |
| read length median± SD | 17,714 ± 2,595 |
| Depth mean ± SD | 32.95 ±4.77 |

##### ONT Nanopore dataset

| | | | RAW Sequencing Results | | | | | |
|---|---|---|---|---|---|---|---|---|
| Participant | Relationship to Proband | Sample Type | Wambam N50 | total.gbp (in Mb) | Coverage (total.gbp / 3.1 Mb) | Median Identity | Number of Flow Cells Used | IDs in Negi et al. 2025 |
| Mother #2 | Self | HMW DNA | 15795 | 58.64563701 | 18.91794742 | 0.9925 | 1 | DSDTRN19 |
| Proband #3 | Self | HMW DNA | 27840 | 78.115 | 25.1983871 | 0.987 | 1 | DSDTRN09 |
| Proband #4 | Self | Whole Blood - 750 -800 μL | 21984 | 133.8740091 | 43.18516422 | 0.9925 | 2 | DSDTRN04 |
| Father #4 | Father | Whole Blood - 750 -800 μL | 32674 | 105.6758067 | 34.08896989 | 0.9915 | 1 | DSDTRN05 |
| Mother #4 | Mother | Whole Blood - 750 -800 μL | 29469 | 84.60645869 | 27.29240603 | 0.9925 | 1 | DSDTRN06 |
| Proband #5 | Self | Whole Blood - 750 -800 μL | 29507 | 86.72254049 | 27.97501306 | 0.9915 | 1 | DSDTRN11 |
| Proband #6 | Self | WBCs - 800 μL | 36036 | 130.1191238 | 41.97391089 | 0.9925 | 1 | DSDTRN15 |
| | | | **29585** | **103.1854898** | **33.28564187** | **0.99125** | | |

| Cohort-wide statistics (n=98) | | | | |
|---|---|---|---|---|
| median | 33534 | 112.89 | 36.4 | 0.9915 |
| average | 32127 | 113.86 | 36.7 | 0.9922 |

##### OGM Bionano dataset

| Participant | Enzyme | Site | Total DNA (>= 150kbp) | N50 (>= 150kbp) | Map rate | Effective coverage | Average label density (≥ 150kbp) |
|---|---|---|---|---|---|---|---|
| Pariticpant#1 | DLE-1 | CTTAAG | 1,206.3 Gbp | 202.13 kbp | 80.80% | 306.39 | 15.28 /100kbp |
| Proband #2 | DLE-1 | CTTAAG | 1,128.68 Gbp | 263.63 kbp | 87% | 309.04 | 15.53 /100kbp |
| Father #2 | DLE-1 | CTTAAG | 1,105.11 Gbp | 230.25 kbp | 80.00% | 276.5 | 15.63 /100kbp |
| Mother #2 | DLE-1 | CTTAAG | 1,104.85 Gbp | 222.38 kbp | 77.30% | 268.95 | 15.21 /100kbp |
| Proband #3 | DLE-1 | CTTAAG | 1,034.31 Gbp | 274.88 kbp | 94.20% | 300.58 | 15.27 /100kbp |
| Proband #7 | DLE-1 | CTTAAG | 613.86 Gbp | 204.38 kbp | 68.40% | 129.57 | 13.43 /100kbp |

**Supplementary Fig. S1**

Visualization in IGV of the location of one of the CTTAGG sequences recognized for Optical Genome Mapping DLE1 labeling (marked as an orange triangle in manuscript figures).

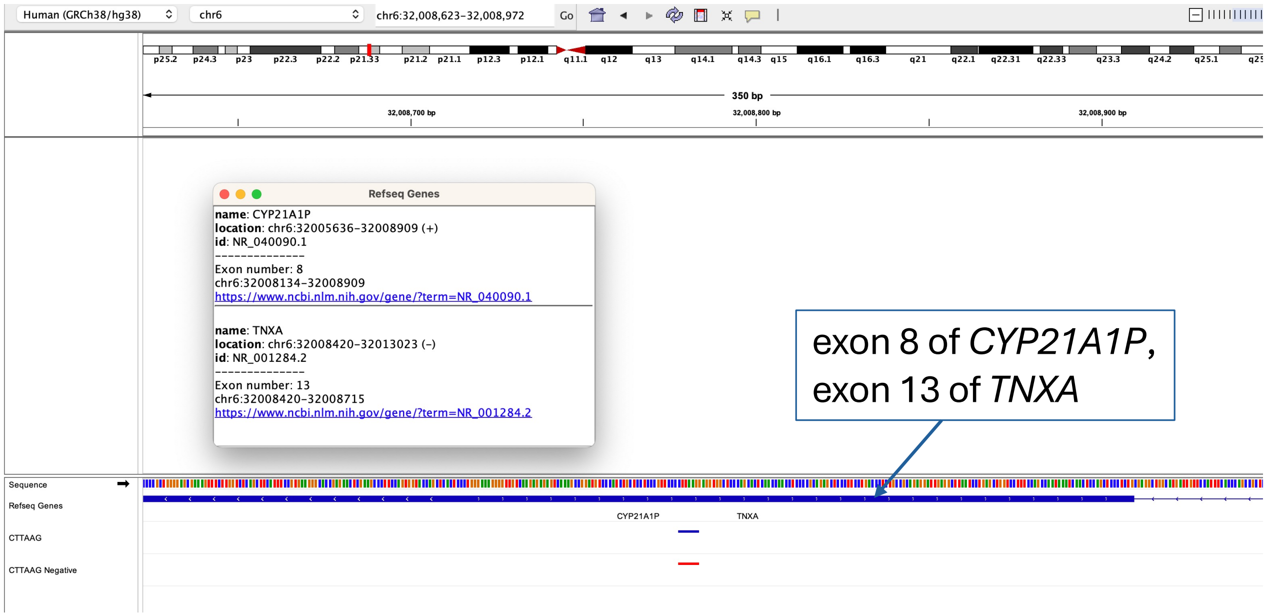

#### Supplementary Fig. S2

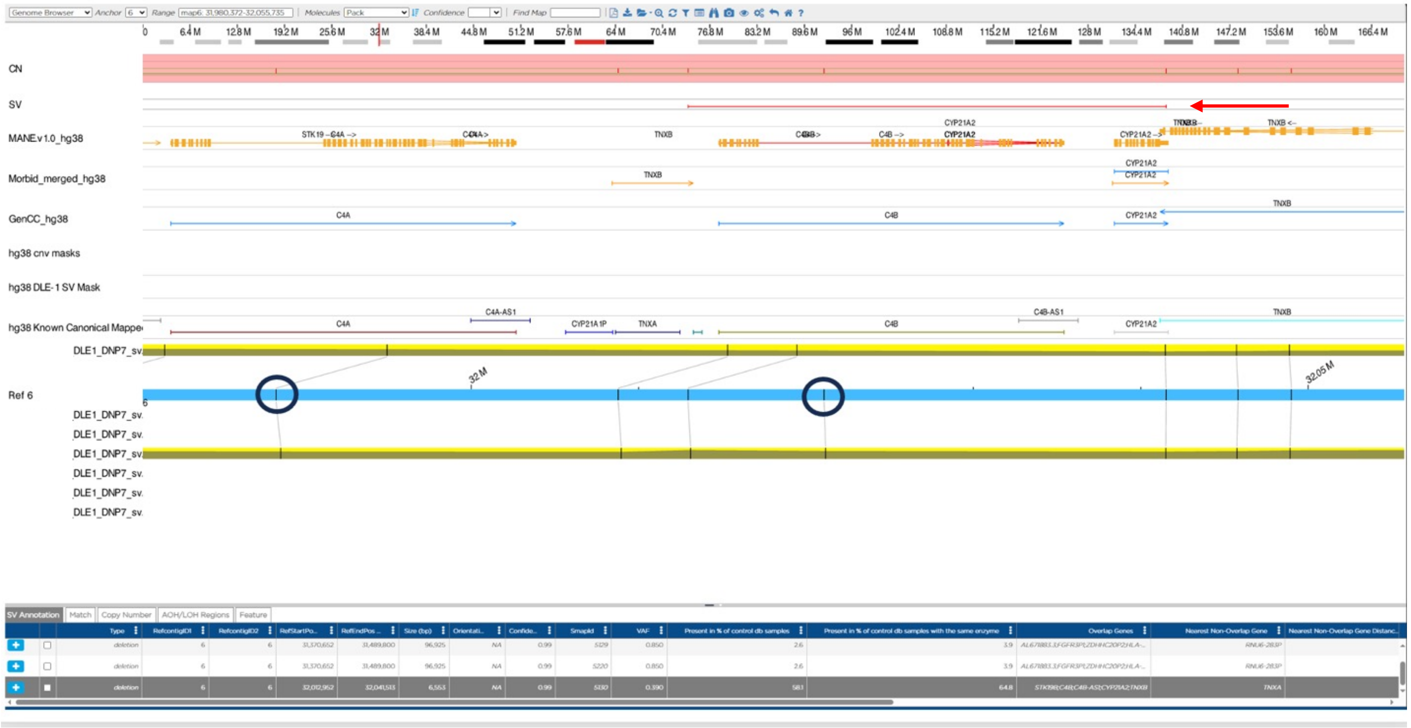


**Optical Genome Mapping on Proband #7**

Bionano Access software output of the DLE-DNP7-sv tool aligning to reference hg38 showed two bimodular alleles (olive green) in the proband. The labels situated in the HERV-K retrovirus are circled over the hg38 reference assembly (blue).

In the top haplotype the 1st RCCX module harbors HERV-K in the *C4* gene, the 2nd module does not. This common polymorphism is accurately called as a 6.5 kb deletion (greyed row in table, red bar in SV track), red arrow. In the bottom haplotype the transposon is present in both modules, as in the hg38 reference.

In both haplotypes, the label at the end of the active *CYP21A2* gene was present and the full gene deletion detected clinically in this patient was not visualized or called.

hg 38 (DLE1 assembly 7)

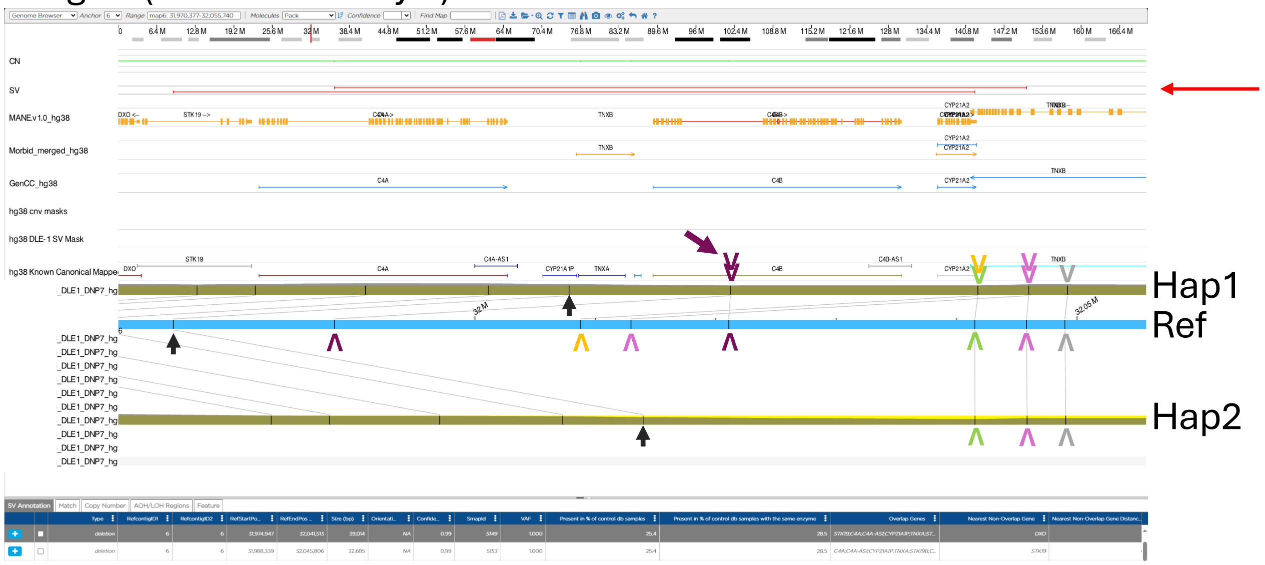


hg 19 (DLE1 assembly 6)

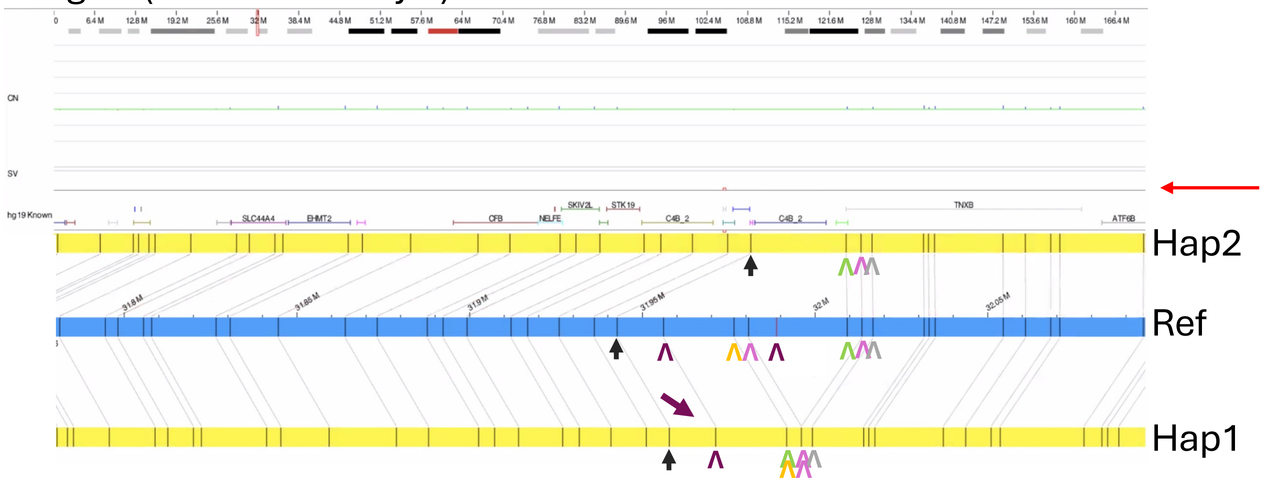


### Supplementary Fig. S3

#### Optical Genome Mapping on Participant #1

The reference assembly is shown in blue in these Access software screenshots. Proband assemblies are shown in olive and yellow in the alignments to hg38 (top) and hg19 (bottom) respectively.

Black arrows indicate the first label in the RCCX module, in *STK19* upstream of *C4*. Other RCCX labels are highlighted as follows:

- V HERV-K in *C4*
- V *CYP21A2*
- V *CYP21A1P*
- V *TNXA* or *TNXB*
- V Distal TNXB-specific

In Haplotype 1, both TNXA/B (pink) labels of the reference map to a single label in the proband. Similarly, the *CYP21A1P* and *CYP21A2* reference labels map to a single label. This points to a monomodular haplotype (with HERV-K) carrying either the gene or the pseudogene. A 32,685 bp deletion is called. (Metrics are shown in the table below the assemblies.)
In the second haplotype, in the complete absence of HERV-K, the software calls a 39 kb, deletion and aligns the single CYP21 label to the active gene, suggesting a deletion of the pseudogene.

Earlier analysis using hg19 as the reference and a previous version of the assembly tool had yielded a slightly different alignment for Hap1 where the single HERV-K label in the participant mapped to only the "1st module" position of the reference, not both (Purple arrow ↘). And the Access software did not call the deletions (see red lines in SV track up the upper panel, red arrows).
Clinical testing identified only one pseudogene and one gene in this unaffected sibling. Hap1 may be the deletion allele (1 pseudogene, no gene), and Hap2 may be a monomodular allele with only the intact gene and no module carrying the pseudogene.

### Supplementary Fig. S4

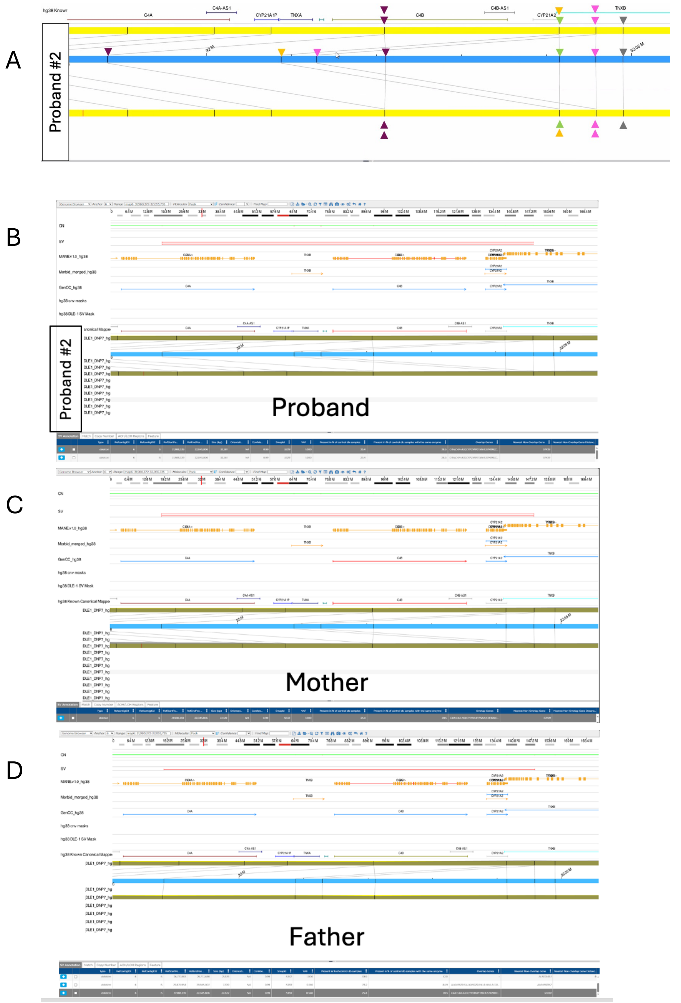


**Optical Genome Mapping on Trio #2**

Access software output shown for DLE1_DNP7_hg38. Expected alleles in the proband (A, B) include one with Int2G variant and one allele with full gene deletion. While clinical testing was performed at Mayo Genetics Laboratories, the original report is no longer available to the current clinical team and information about the number of pseudogenes has been lost.

Here the two haplotypes were indistinguishable and appeared identical to the haplotype we termed Hap2 found Proband #3 and Participant #1 (see Fig. 3). This appears to be a monomodular allele, with each label of the RCCX module mapped by the Access software to two positions on the reference assembly (color code for triangles as in Fig. 3A). A 32,581 kb deletion is called on each. Whether the pseudogene or the active *CYP21A2* is missing could not be determined.

The unaffected mother (C) also appeared homozygous for Hap2 with a 32,519 bp deletion called. The father (D) appeared heterozygous with one haplotype similar to the proband's and mother's, except with a slightly larger (32,622 bp) deletion, and one that aligns directly to the reference (bimodular with HERV-K in each copy of the *C4* gene).

### Supplementary Fig. S5: Optical Genome Mapping Proband #3

The only difference noted between the hg19 (top panel) and hg38 alignments in this proband was that the two labels common to *TNXA* and *TNXB* (pink triangles) did not comap on hg19 (pink arrow). 6.5 kb and ~32.5 kb deletions called on both references (Red arrow, top panel; see also Fig. 3A).

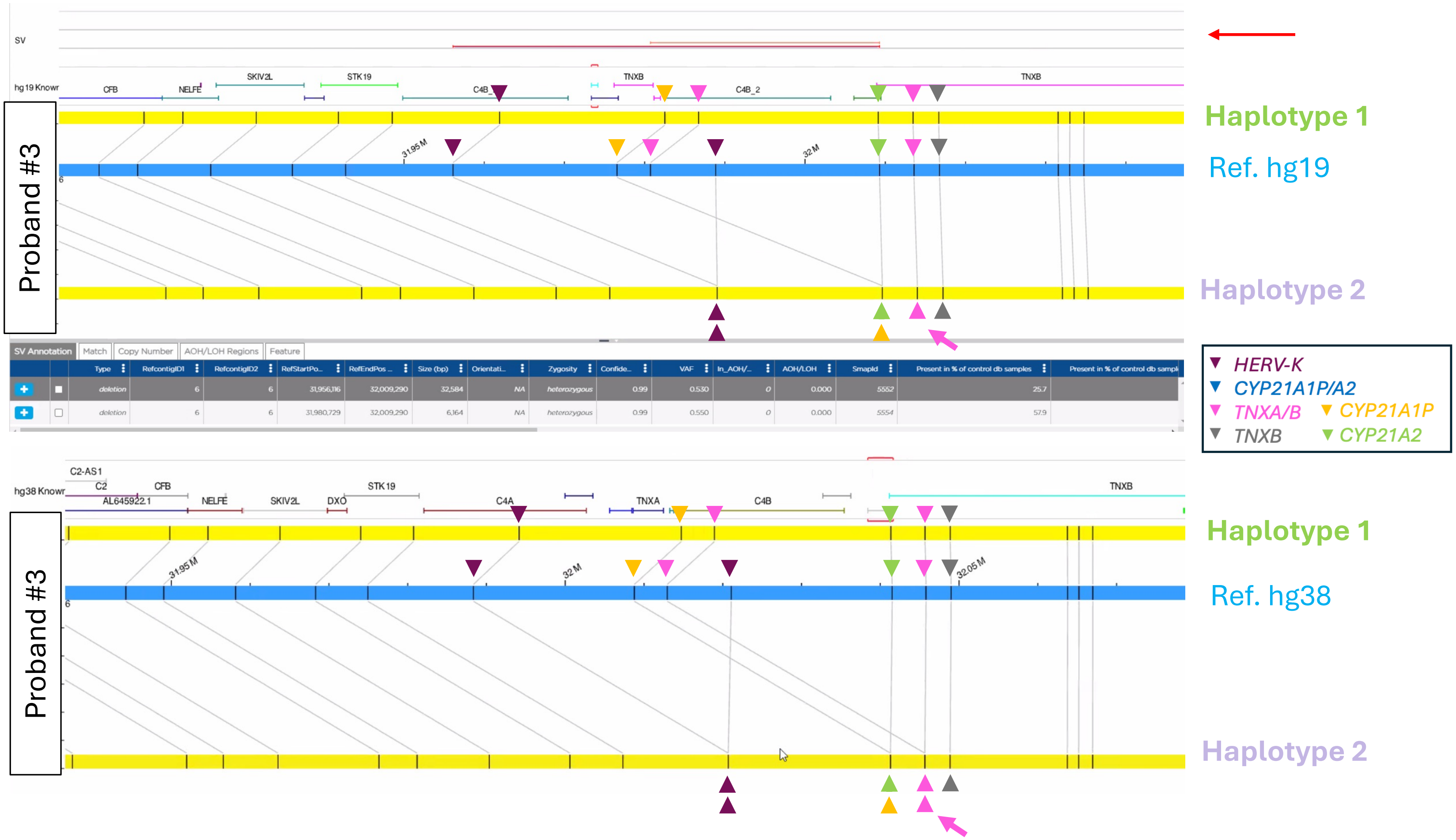

#### Supplementary Fig. S6

***Parakit* output for Trio #4 (nanopore-based LRS)**

The Parakit output of the *Variant-Finder* tool (top panel) showed reads supporting a fusion in intron 3 of *CYP21A2* (triangle) and a Q319X SNV (circle). [Note that the read ID number provided in the raw output was replaced here with "supp_read" for figure clarity.]

*Allele-Support* (aggregated coverage and module 2 ratio), and *Haplotype-Finder* (predicted haplotypes) tools are shown for Proband #4 and her parents.

In the proband, module 2 ratio showed a clear shift at the site of the predicted intragenic fusion, while coverage remained stable around 3. The fusion was inherited from the mother (H1 haplotype in the proband, highlighted in blue). The Q319X SNV was inherited from the father (proband H2 haplotype, highlighted in red).

The bottom track displays the annotation for RCCX module *CA*, *TNX*, and *CYP21* genes and pseudogenes.

.

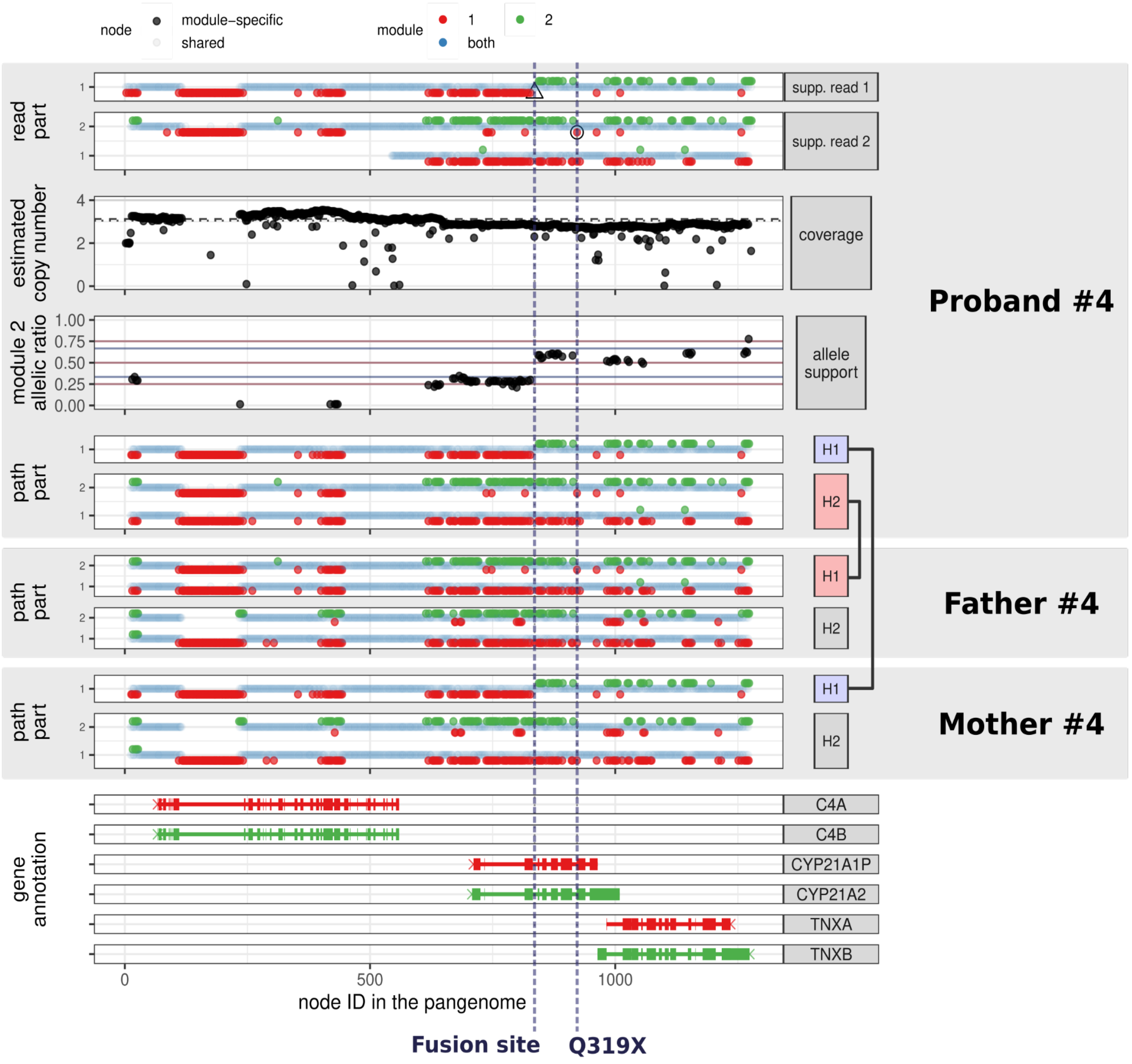

#### Supplementary Fig. S7

##### ***Parakit*** **output for Proband #3 (nanopore-based LRS)**

The output of the *Variant-Finder* tool (top panel showed reads supporting a fusion located beyond the *CYP21A2* gene in *TNXB* (black triangle) and Int2G SNV (black circle). Locations are highlighted by the vertical dotted lines through all panels. *Allele-Support* (aggregated coverage and module 2 ratio), and *Haplotype-Finder* (predicted haplotypes) tool are shown below. The bottom track displays the annotation for RCCX module *CA*, *TNX*, and *CYP21* genes and pseudogenes.

Each predicted haplotype carried one of the variants found by the *Variant-Finder*. Coverage for the region was estimated to be around 3 and a switch in allele support coincided with the predicted fusion site located beyond the *CYP21A2* gene, in the *TNXB* gene, further supporting a complete gene deletion allele.

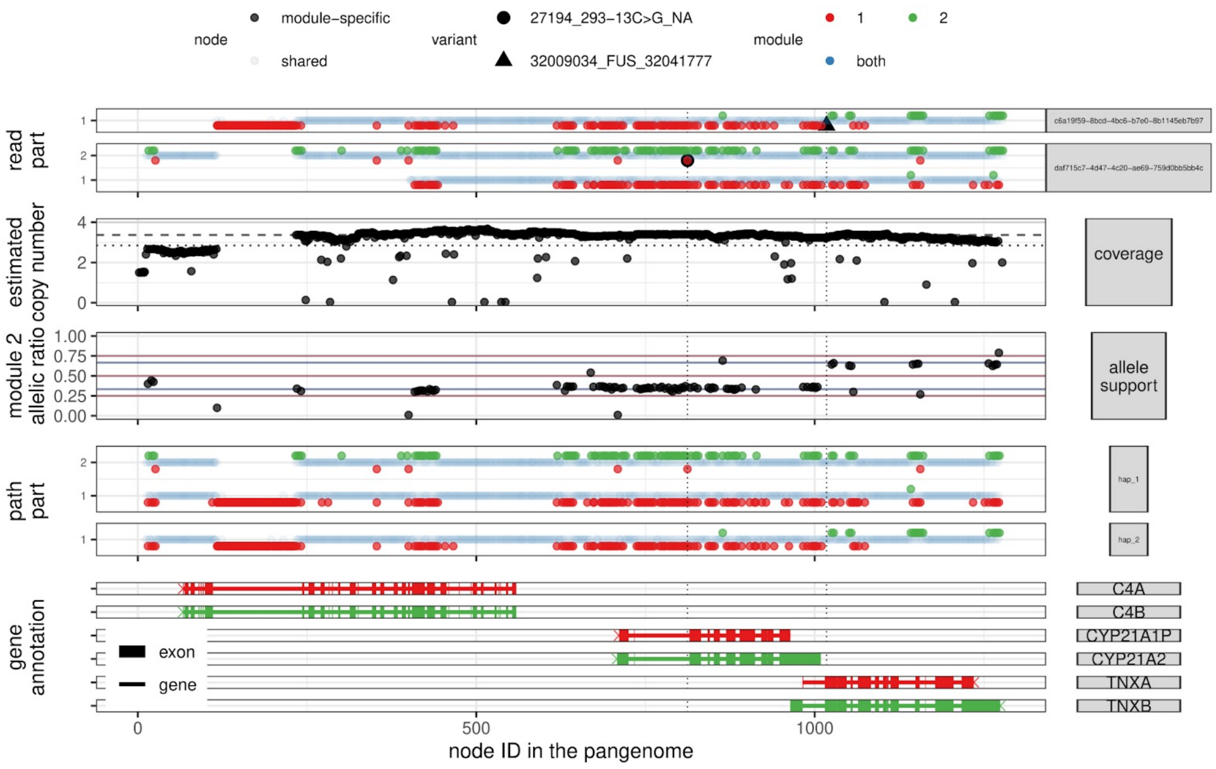

### Supplementary Fig. S8

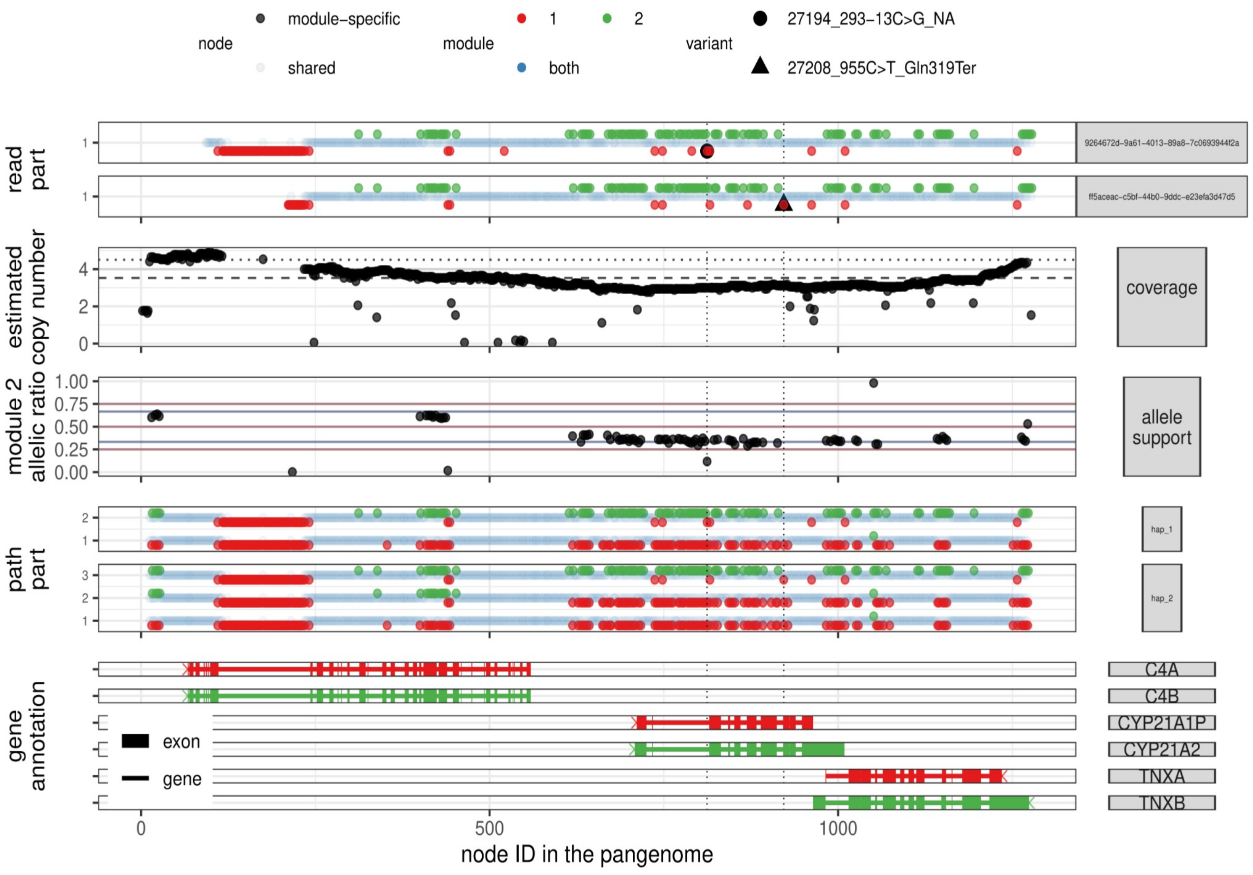


#### ***Parakit*** **output for Proband #5 (nanopore-based LRS)**

The output of the *Variant-Finder* tool showed reads supporting Q319X (black triangle) and Int2G (black circle) SNVs in compound heterozygous configuration (top panel). *Allele-Support* (aggregated coverage and module 2 ratio), and *Haplotype-Finder* (predicted haplotypes) tool are shown below. The bottom track displays the annotation for RCCX module *CA*, *TNX*, and *CYP21* genes and pseudogenes.

Each predicted haplotype carried one of the variants found by the *Variant-Finder* (see longest supporting reads in the top panel) highlighted by the vertical dotted guides. Coverage showed signs of bias as it was above 4 copies at the flanks and gradually decreased down to 3 in the middle of the module. This bias may explain why even at the flanks copy number is underestimated (~4.5 instead of the expected 5 based on the predicted haplotypes). Of note, we observed signs of gene conversion between *C4A* and *C4B* in the second module of haplotype Hap2, which also may affect coverage calculation. Predicted Hap2 was CA4-CYP21A1P-TNXA **C4B**-CYP21A1P-TNXA C4B-CYP21A2-TNXB, with *C4B* instead of *C4A* in the middle module of this tri-modular haplotype (see “green” dots in Path Part 2 of Hap 2).

#### Supplementary Fig. S9

***Parakit* output for Mother #2 (nanopore-based LRS)**

In this unaffected mother, the *Variant-Finder* tool identified only reads supporting one pathogenic allele (vs. two in each of the affected probands; black circle), a fusion toward the end of the *CYP21A2* active gene. In this low-quality sample, the longest read (top panel) did not extend over the entirety of the *C4* gene. Estimated copy number (aggregated coverage panel) was consistently around 2 suggesting two monomodular haplotypes. Module 2 ratio was at 0.5 and increased after the predicted fusion. Predicted haplotypes are monomodular, with a shift from red (pseudogene-type module) to green, within an imprecise window. Note the absence of informative data around the predicted fusion, with the last "red" pangenome node present around the penultimate exon of *CYP21A2*, and the first "green" node in the *TNXB* gene (highlighted with a wider pink arrow in Fig. 6.)

The bottom track displays the annotation for RCCX module *CA*, *TNX*, and *CYP21* genes and pseudogenes.

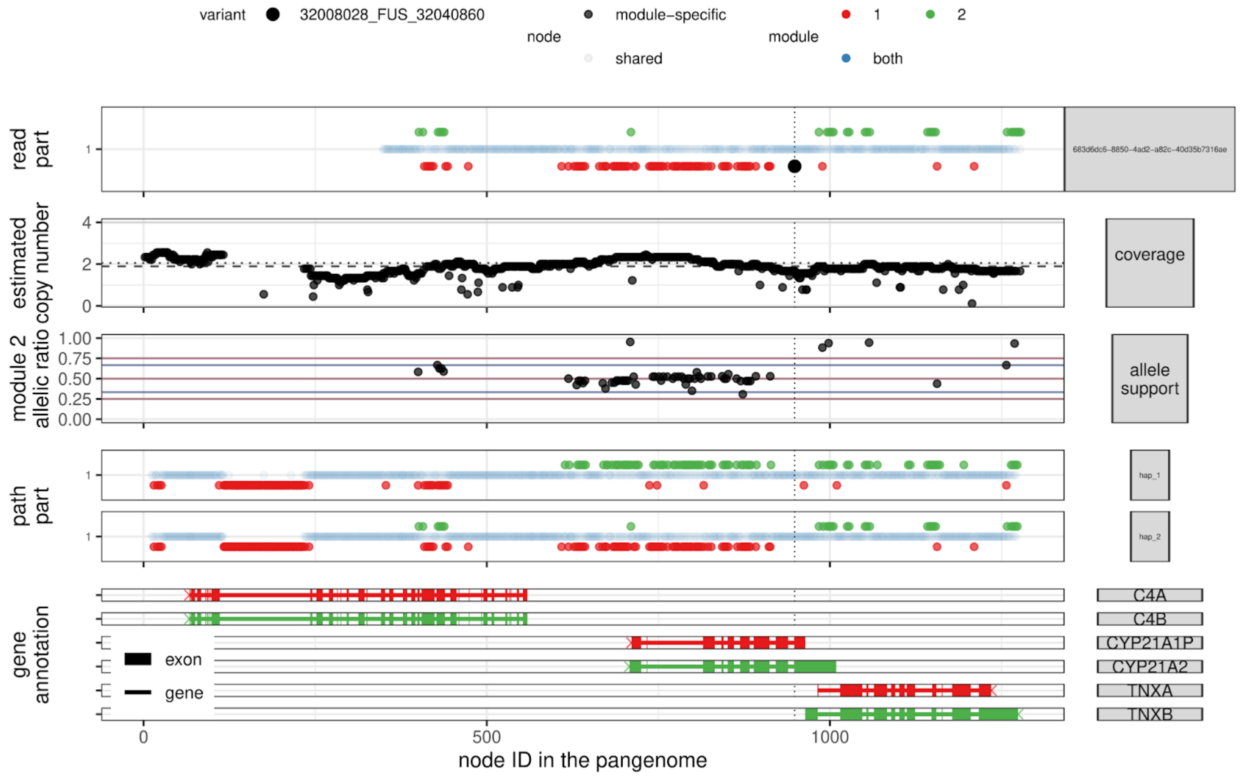
